# 2025/26 subclade-specific influenza A and B vaccine effectiveness estimates and discordant laboratory-based indicators of vaccine match or mismatch

**DOI:** 10.64898/2026.09.10.26362769

**Authors:** Lea Separovic, Samantha E Kaweski, Michelle B Cox, Yuping Zhan, Suzana Sabaiduc, Romy Olsha, Maan Hasso, Richard G Mather, Sara Carazo, Christine Lacroix, Isabelle Meunier, Lila N Salhi, James A Dickinson, Nathan Zelyas, Agatha N Jassem, Katie Dover, Charlene Ranadheera, Ruimin Gao, Nathalie Bastien, Danuta M Skowronski

## Abstract

**Background:** The 2025/26 influenza season included A(H3N2) subclade K predominance with A(H1N1)pdm09 co-circulation and subsequent prolonged influenza B activity. The Canadian Sentinel Practitioner Surveillance Network reports vaccine effectiveness (VE) in the context of genetic and antigenic relatedness to circulating viruses.

**Methods:** We estimated VE by test-negative design among outpatients with medically-attended acute respiratory illness between November and April. We characterized case viruses by whole genome sequencing and hemagglutination inhibition assay.

**Results:** Among 2809/9047 (31%) influenza-positive specimens, 82% (2306/2809) were influenza A with 18% (504/2809) influenza B. Of subtyped influenza A, 85% (1848/2169) were A(H3N2).

We sequenced 59% (1099/1848) of A(H3N2) viruses, with 87% (956/1099) subclade K, antigenically-mismatched to subclade J.2 vaccine. VE against A(H3N2) and subclade K was 38% (95% confidence interval: 27%-47%) and 33% (19%-45%).

We sequenced 69% (222/323) of A(H1N1)pdm09 viruses, virtually all (n=218) clade 5a.2a.1, including subclades D.3.1 (21%; n=47) and D.3.1.1 (77%; n=171), antigenically-matched to subclade D vaccine. VE against A(H1N1)pdm09, subclades D.3.1 and D.3.1.1 was 28% (4%-47%), −21% (−144%-40%) and 44% (14%-64%).

We sequenced 82% (413/504) of influenza B viruses, all B/Victoria, including antigenically-mismatched subclade C.3 (18%; n=74) with D197N gain-of-glycosylation, and antigenically-matched subclade C.5 (82%; n=339) relative to subclade C vaccine. VE against influenza B, subclades C.3 and C.5 was 55% (41%-66%), 10% (−60%-49%) and 68% (51%-79%).

**Conclusions:** The 2025/26 influenza vaccine reduced the risk of A(H3N2), A(H1N1)pdm09 and B/Victoria by over one-third, one-quarter and one-half, respectively. Laboratory-based indicators of vaccine match remain uncertain proxies for actual vaccine protection, reinforcing importance of annual epidemiological monitoring of VE.

## Introduction

The 2025/26 influenza season in Canada, as elsewhere in the northern hemisphere (NH), was marked by predominant influenza A(H3N2) circulation, notably the emergent subclade K variant [1–6]. Early laboratory-based surveillance revealed multiple mutations at key antigenic sites of the hemagglutinin (HA) glycoprotein of subclade K, contributing to substantial antigenic drift and raising widespread concern for reduced vaccine protection. The rest of the influenza season was characterized by influenza A(H1N1)pdm09 co-circulation and a subsequent prolonged influenza B epidemic [3,6], with a mix of genetic variants contributing.

Conventional laboratory indicators of vaccine match or mismatch, both genotypic and phenotypic, are needed for timely detection of emerging variants potentially warranting vaccine strain update. In vitro indicators, however, remain uncertain intermediaries for predicting actual vaccine protection. Epidemiological monitoring of vaccine effectiveness (VE) is ultimately required to quantify the relative reduction in risk conferred among vaccinated versus unvaccinated people. In that regard, and notwithstanding laboratory indicators of vaccine mismatch, mid-season estimates of 2025/26 influenza VE against subclade K were within historic expectation for A(H3N2) [7–10]. For highly changeable viruses such as influenza, an integrated package of genetic, antigenic and epidemiologic evidence is needed to inform annual vaccine performance and its multi-factorial virological and immuno-epidemiological determinants.

Since the 2005/06 season, the Canadian Sentinel Practitioner Surveillance Network (SPSN) has directly incorporated genetic and antigenic characterization of contributing case viruses in annual influenza VE estimation, undertaken for context, cross-validation and clade-specific estimation as ultimately required to inform vaccine strain selection [11,12]. Here, we report full 2025/26 end-of-season VE estimates including subclade-specific A(H3N2), A(H1N1)pdm09, and B/Victoria findings.

## Methods

### Canadian SPSN

The Canadian SPSN includes community-based sentinel practitioners in Canada’s four largest provinces: Alberta, British Columbia, Ontario, and Quebec. Sentinels collect respiratory specimens from consenting patients presenting with acute respiratory illness (ARI; defined as new or worsening cough potentially due to infection) within 7 days of illness onset. Accredited provincial laboratories test specimens with real-time RT-PCR and/or multiplex assays. Current analyses include participants ≥1 year old with specimens collected between epi-week 44–17 (26 October 2025 to 2 May 2026).

### Influenza vaccines

Publicly-funded trivalent influenza vaccine administration began in October in SPSN provinces, primarily using inactivated (≥99%) and egg-based (≥90%) products. Adjuvanted vaccine was offered to community-dwelling older adults ≥65 years (≥75 years in Quebec); in Ontario, high-dose vaccine was more often available but without preferential recommendation over adjuvanted vaccine. Only the A(H3N2) vaccine strain was updated, from clade 2a.3a.1 subclade J in 2024/25 to subclade J.2 in 2025/26 [13]. The A(H1N1)pdm09 vaccine strain was unchanged, remaining clade 5a.2a.1 subclade D (egg-based) or C.1.1 (cell-based) since 2023/24 [13]. The influenza B/Victoria vaccine strain was also unchanged, remaining clade V1A.3a.2 subclade C since 2022/23 [13].

### Epidemiologic analyses

Logistic regression was used to estimate odds ratios (ORs) comparing influenza positivity between vaccinated and unvaccinated participants, adjusting for age group, province, and calendar time, with VE calculated as (1-OR)*100%. Sensitivity analyses explored additional adjustment for sex and comorbidity and the impact of excluding SARS-CoV-2-positive specimens from influenza controls due to potentially-correlated vaccination behaviours [14,15]. Firth’s regression addressed sparse data issues [16]. To explore repeat vaccination effects, we estimated VE for those with current-season-only, current-and-prior-season, or prior-season-only vaccination, compared to those with neither-season vaccination.

### Virologic characterization

We attempted whole genome sequencing of eligible case viruses following previously described methods at both provincial and national laboratories [7]. HA clades and subclades were assigned as per Nextclade [17], specifying amino acid substitutions and affected antigenic sites in parentheses relative to cell-based reference strains, additionally annotating involvement of the receptor binding site (RBS) and gain/loss of glycosylation (+/-CHO) (**Supplementary Tables S1-S2**).

We antigenically characterized a subset of cell-propagated SPSN case viruses by hemagglutination inhibition (HI) assay. By convention we defined antigenic distinction (vaccine mismatch) as ≥8-fold reduction in HI titre against the case virus relative to the HI titre against the comparator vaccine reference strain. Our primary approach used ferret antisera raised against cell-based vaccine antigen with comparator titre measured against the homologous cell-based vaccine reference virus. Given foremost use of egg-based vaccines, we also undertook sensitivity HI characterization using egg-based vaccine ferret antisera with comparator titre measured against the homologous egg-based vaccine reference virus. Lastly, to better mimic egg-based vaccine-induced responses to cell-propagated viruses (as per SPSN case viruses), we explored using egg-based vaccine ferret antisera with the heterologous cell-based vaccine reference titer as the comparator (**Supplementary Methods**).

## Results

### Influenza detections

Among 9047 included specimens, 31% (2809) were influenza positive with 82% (2306) influenza A and 18% (504) influenza B (including one influenza A+B co-infection) (**Figure 1**). Of 2169 (94%) subtyped influenza A viruses, 1848 (85%) were A(H3N2) and 323 (15%) were A(H1N1)pdm09 (including one A(H3N2)+B and two A(H3N2)+A(H1N1)pdm09 co-infections). Influenza A(H3N2) positivity peaked at 51% in epi-week 51, followed by later increase in influenza B positivity that remained ≥15% from epi-weeks 9-15.

**Figure 1.**
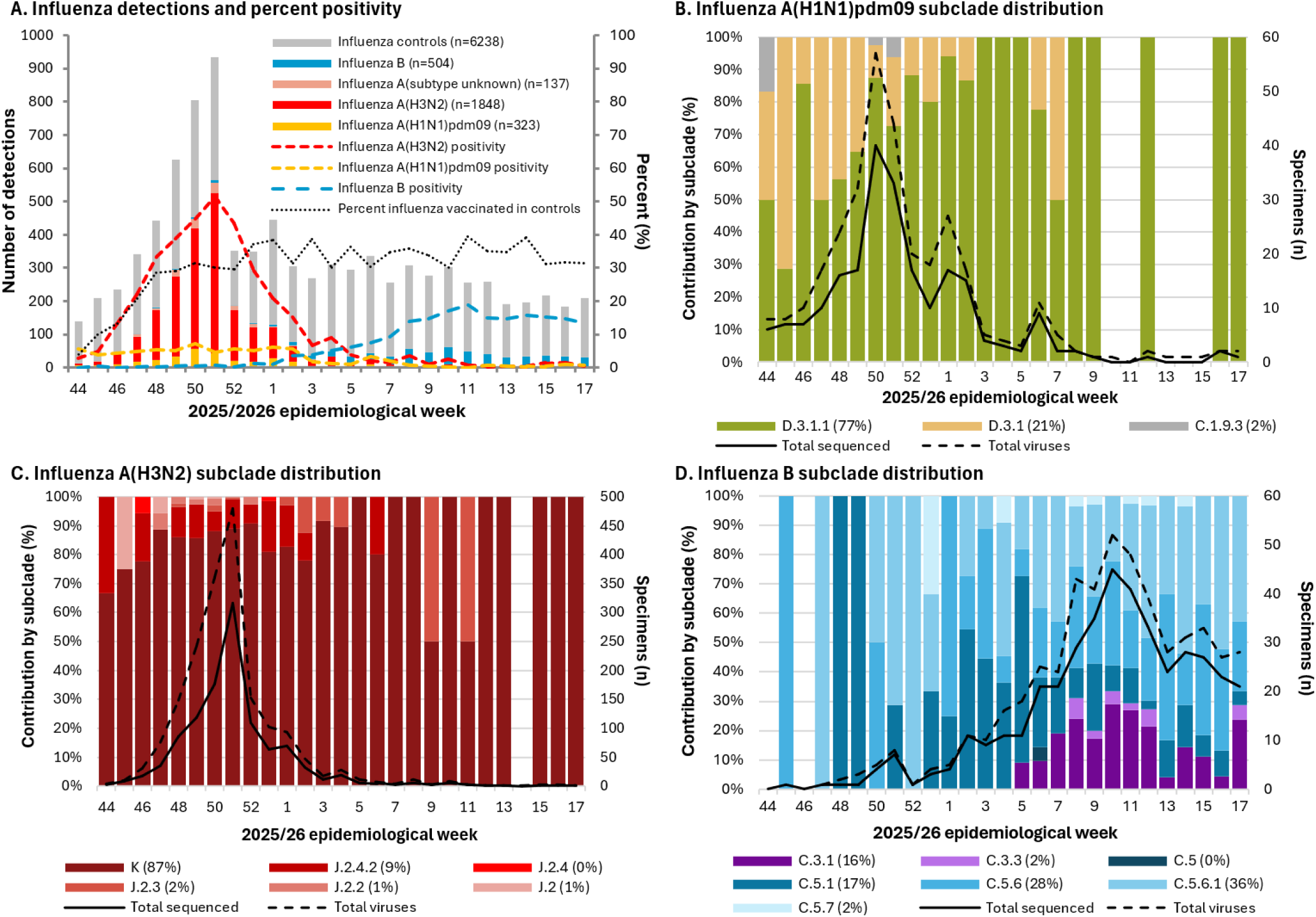
Influenza test-positive and test-negative specimens, including genetic characterization, by week of specimen collection, Canadian Sentinel Practitioner Surveillance Network, 26 October 2025 – 02 May 2026 (weeks 44–17) (n=9050). Influenza test-positive and test-negative specimen tallies by epi-week are shown in panel A with overlaid percent positivity (dashed lines) and percent vaccinated ≥2 weeks before onset (dotted line). Subclade distributions, total number of specimens (solid line), and total number of sequenced specimens (dashed line) for those contributing to vaccine effectiveness analyses are shown by epi-week in panel B for influenza A(H1N1)pdm09, panel C for influenza A(H3N2), and panel D for influenza B. Displayed detections in panel A include one influenza A(H3N2) and B co-infection and two influenza A(H1N1)pdm09 and A(H3N2) co-infections. These six viruses from three specimens are displayed separately, thus totaling n=9050 specimens in panel A (n=9047 when co-infections not counted separately). All A(H3N2) subclades displayed in panel C belong to clade 2a.3a.1. Among displayed A(H1N1)pdm09 subclades in panel B, all belong to clade 5a.2a.1 except C.1.9.3 (n=4), belonging to clade 5a.2a. All influenza B subclades in panel D belong to clade V1A.3a.2 For further details on virological characterization, subclade designations, and tallies by province, see **Supplementary Tables S1 and S2**.

### Virologic characterization

We sequenced 59% (1099/1848) of A(H3N2) case viruses (**Supplementary Table S1**). All belonged to clade 2a.3a.1 but unlike the subclade J.2 vaccine, 87% (956/1099) were subclade K. Overall, most (96%; 1059/1099) sequenced viruses bore subclade J.2.4 clade-defining substitutions T135K (A)(RBS)(− CHO) and K189R (B), with additional S144N (A)(+CHO), N158D (B), I160K (B) and T328A mutations, common to descendant subclades K (formerly J.2.4.1) and J.2.4.2. Of these, 90% (956/1059) were subclade K, characterized by additional K2N, Q173R (D) and HA2:S49N; the remaining 10% (103/1059) were subclade J.2.4.2, characterized by F79V. Among 157 antigenically-characterized A(H3N2) viruses, all subclade K, J.2.4.2, and J.2.3 viruses were vaccine-mismatched, whereas all J.2 and J.2.2 viruses were vaccine-matched (**Table 2**). Among viruses showing antigenic distinction, fold-reductions were greater in relation to egg-based vaccine reference virus and antisera (**Supplementary Table S3**).

We sequenced 69% (222/323) of A(H1N1)pdm09 case viruses (**Supplementary Table S1**). Almost all (98%; 218) belonged to clade 5a.2a.1, but unlike subclade D vaccine, 47 (21%) were subclade D.3.1, distinguished by T120A and HA2 substitutions I45V, I133T and V193A. Increasing across the season (**Figure 1**), 171 (77%) were subclade D.3.1.1 defined by R113K, A139D (Ca2), E283K, and K302E substitutions with 15% (25/171) further distinguished by D139N (Ca2) evolution. Among 81 antigenically-characterized A(H1N1)pdm09 viruses, all but one (D.3.1) were vaccine-matched (**Table 2**), similar in relation to egg-based vaccine reference virus and antisera (**Supplementary Table S4**).

We sequenced 82% (413/504) of influenza B case viruses (**Supplementary Table S2**). All were influenza B/Victoria belonging to clade V1A.3a.2 but unlike the subclade C vaccine, 18% (74) were subclade C.3, distinguished by E128K (120-loop) and A154E, and 82% (339) were subclade C.5, distinguished by D197E (190-helix)(RBS). The C.3 viruses all had additional D197N (190-helix)(RBS)(+CHO), defining for both circulating subclades C.3.1 (81%; 60/74) and C.3.3 (19%; 14/74), the latter further characterized by S208P, S255P and I267V. The C.5 viruses were most commonly subclades C.5.6 (34%; 114/339), defined by D129N (120-loop), and C.5.6.1 (44%; 148/339), further defined by T37I, E128D (120-loop) and T199A (190-helix)(RBS). Among 97 antigenically-characterized B/Victoria viruses, all C.3 were vaccine-mismatched whereas all C.5 were vaccine-matched. In sensitivity HI assays using egg-raised antisera and homologous egg-based vaccine comparator, all influenza B viruses were considered vaccine-mismatched, but with C.3 viruses still showing greater fold-reductions than C.5 (≥128-fold vs. 16-32-fold, respectively). Using egg-raised antisera but instead heterologous cell-based vaccine comparator, findings reverted to primary homologous cell-based observations indicating C.5 viruses were vaccine-matched (**Supplementary Table S5**).

### Participant characteristics

Participant profiles for those included in influenza A(H3N2) analyses are displayed in **Table 1** and for influenza A(H1N1)pdm09 and B in **Supplementary Tables S6-S7**. Among SPSN controls ≥18 years, the proportion vaccinated with (34%) or without (36%) regard to timing was comparable to 2025/26 Canadian coverage survey estimates among ≥18-year-olds (34%)[18], reassuring against systematic sampling bias. As previously found [14], about half of influenza-vaccinated controls also self-reported 2025/26 COVID-19 vaccine receipt, increasing with age (**Supplementary Table S8)**.

**Table 1.** Participant profile, influenza A(H3N2) analyses, Canadian Sentinel Practitioner Surveillance Network, 26 October 2025 – 02 May 2026 (epi-weeks 44–17) (n=8086).

| Characteristics | All ARI participants (column %) |  |  |  |  |  | Influenza vaccinated<sup>a</sup> (row %) |  |  |  |  |  |
| --- | --- | --- | --- | --- | --- | --- | --- | --- | --- | --- | --- | --- |
|  | Overall |  | Influenza A(H3N2) cases |  | Influenza controls |  | Overall |  | Influenza A(H3N2) cases |  | Influenza controls |  |
|  | n | % | n | % | n | % | n | % | n | % | n | % |
| N (row %) | 8086 | 100 | 1848 | 23 | 6238 | 77 | 2261 | 28 | 340 | 18 | 1921 | 31 |
| **Age group (years) <sup>b</sup>** |  |  |  |  |  |  |  |  |  |  |  |  |
| 1–8 | 1283 | 16 | 429 | 23 | 854 | 14 | 238 | 19 | 58 | 14 | 180 | 21 |
| 9–17 | 885 | 11 | 382 | 21 | 503 | 8 | 115 | 13 | 39 | 10 | 76 | 15 |
| 18–49 | 2973 | 37 | 620 | 34 | 2353 | 38 | 598 | 20 | 85 | 14 | 513 | 22 |
| 50–64 | 1374 | 17 | 190 | 10 | 1184 | 19 | 404 | 29 | 35 | 18 | 369 | 31 |
| ≥ 65 | 1571 | 19 | 227 | 12 | 1344 | 22 | 906 | 58 | 123 | 54 | 783 | 58 |
| Median (IQR) | 38 (16–60) |  | 24 (9.5–46) |  | 42 (21–62) |  | 58 (35–72) |  | 44 (15–71.5) |  | 59 (36–72) |  |
| **Sex** |  |  |  |  |  |  |  |  |  |  |  |  |
| Female | 4842 | 60 | 997 | 54 | 3845 | 62 | 1413 | 29 | 187 | 19 | 1226 | 32 |
| Male | 3212 | 40 | 845 | 46 | 2367 | 38 | 846 | 26 | 152 | 18 | 694 | 29 |
| Unknown | 32 | 0 | 6 | 0 | 26 | 0 | 2 | 6 | 1 | 17 | 1 | 4 |
| **Comorbidity<sup>c</sup>** |  |  |  |  |  |  |  |  |  |  |  |  |
| No | 5838 | 72 | 1452 | 79 | 4386 | 70 | 1259 | 22 | 210 | 14 | 1049 | 24 |
| Yes | 1790 | 22 | 304 | 16 | 1486 | 24 | 831 | 46 | 104 | 34 | 727 | 49 |
| Unknown | 458 | 6 | 92 | 5 | 366 | 6 | 171 | 37 | 26 | 28 | 145 | 40 |
| **Province** |  |  |  |  |  |  |  |  |  |  |  |  |
| Alberta | 1110 | 14 | 247 | 13 | 863 | 14 | 342 | 31 | 46 | 19 | 296 | 34 |
| British Columbia | 1430 | 18 | 225 | 12 | 1205 | 19 | 549 | 38 | 48 | 21 | 501 | 42 |
| Ontario | 3881 | 48 | 1069 | 58 | 2812 | 45 | 1130 | 29 | 215 | 20 | 915 | 33 |
| Quebec | 1665 | 21 | 307 | 17 | 1358 | 22 | 240 | 14 | 31 | 10 | 209 | 15 |
| **Weeks of specimen collection, 2025/26<sup>d</sup>** |  |  |  |  |  |  |  |  |  |  |  |  |
| 44–47 | 865 | 11 | 120 | 6 | 745 | 12 | 108 | 12 | 9 | 8 | 99 | 13 |
| 48–51 | 2545 | 31 | 1231 | 67 | 1314 | 21 | 613 | 24 | 221 | 18 | 392 | 30 |
| 52–2<sup>e</sup> | 1321 | 16 | 395 | 21 | 926 | 15 | 409 | 31 | 87 | 22 | 322 | 35 |
| 3–6 | 1111 | 14 | 64 | 3 | 1047 | 17 | 365 | 33 | 13 | 20 | 352 | 34 |
| 7–10 | 975 | 12 | 26 | 1 | 949 | 15 | 326 | 33 | 7 | 27 | 319 | 34 |
| 11–14 | 752 | 9 | 5 | 0 | 747 | 12 | 278 | 37 | 1 | 20 | 277 | 37 |
| 15–17 | 517 | 6 | 7 | 0 | 510 | 8 | 162 | 31 | 2 | 29 | 160 | 31 |
| **2024/25 influenza vaccination status<sup>f</sup>** |  |  |  |  |  |  |  |  |  |  |  |  |
| No | 4508 | 56 | 1097 | 59 | 3411 | 55 | 265 | 6 | 39 | 4 | 226 | 7 |
| Yes | 2557 | 32 | 505 | 27 | 2052 | 33 | 1673 | 65 | 244 | 48 | 1429 | 70 |
| Unknown | 1021 | 13 | 246 | 13 | 775 | 12 | 323 | 32 | 57 | 23 | 266 | 34 |

Abbreviations: ARI, acute respiratory illness; IQR, interquartile range.
<sup>a</sup> 2025/26 vaccination status based on participant or guardian report. Participants vaccinated <2 weeks before onset of symptoms or with unknown vaccination status or timing were excluded. Only trivalent formulations were used in Canada with virtually all publicly-funded vaccines in SPSN provinces being inactivated (≥99% overall; <5% live-attenuated in BC and Quebec) and egg-based (≥90% overall; <30% cell-based in Alberta and <5% in Ontario). In all provinces, adjuvanted vaccines were available for community-dwelling older adults (≥65 years, ≥75 years in Quebec), with high dose vaccines also offered in Ontario.
<sup>b</sup> Children < 1 year excluded based on variability and/or uncertainty in their age-related vaccine eligibility over the course of the epidemic. Older age strata defined as per usual SPSN analyses predicated upon higher likelihood of chronic comorbidity at ≥ 50 years and higher age-associated risk among adults ≥ 65 years [40].
<sup>c</sup> Includes chronic comorbidities that place individuals at higher risk of serious complications from influenza as defined by Canada’s National Advisory Committee on Immunization [40].
<sup>d</sup> Missing specimen collection dates were imputed as the date the specimen was received and processed at the laboratory minus 2 days.
<sup>e</sup> Includes epi-weeks 52, 53, 1, and 2.
<sup>f</sup> 2024/25 vaccination status based on participant or guardian report.

**Table 2.** Antigenic characterization of a subset of influenza A(H3N2) viruses included in vaccine effectiveness analysis, Canadian Practitioner Surveillance Network, 26 October 2025 – 02 May 2026 (epi-weeks 44–17) (n=335).

|  |  | Antigenically similar or distinct (n) |  | Fold-difference among antigenically distinct (n) |  |  |  |
| --- | --- | --- | --- | --- | --- | --- | --- |
| Subclade | Total tested (n) | Similar | Distinct | 8 | 16 | 32 | ≥64 |
| Influenza A(H3N2) |  |  |  |  |  |  |  |
| J.2 | 4 | 4 | - | - | - | - | - |
| J.2.2 | 5 | 5 | - | - | - | - | - |
| J.2.3 | 7 | - | 7 | - | 5 | 2 | - |
| J.2.4 | 2 | 1 | 1 | 1 | - | - | - |
| J.2.4.2 | 10 | - | 10 | - | 6 | 4 | - |
| K | 107 | - | 107 | 21 | 76 | 10 | - |
| Unknown | 22 | - | 22 | 2 | 15 | 4 | 1 |
| Total | 157 | 10 | 147 | 24 | 102 | 20 | 1 |
| Influenza A(H1N1)pdm09 |  |  |  |  |  |  |  |
| D.3.1 | 30 | 29 | 1 | 1 | - | - | - |
| D.3.1.1 | 42 | 42 | - | - | - | - | - |
| Unknown | 9 | 9 | - | - | - | - | - |
| Total | 81 | 80 | 1 | 1 | - | - | - |
| Influenza B |  |  |  |  |  |  |  |
| C.3.1 | 10 | - | 10 | - | 8 | 2 | - |
| C.3.3 | 5 | - | 5 | 1 | 4 | - | - |
| C.5.1 | 17 | 17 | - | - | - | - | - |
| C.5.6 | 33 | 33 | - | - | - | - | - |
| C.5.6.1 | 24 | 24 | - | - | - | - | - |
| C.5.7 | 4 | 4 | - | - | - | - | - |
| Unknown | 4 | 4 | - | - | - | - | - |
| Total | 97 | 82 | 15 | 1 | 12 | 2 | - |

Hemagglutination inhibition (HI) assays were performed using the 2025/26 recommended **cell-based vaccine strains** for influenza A(H3N2) (A/District of Columbia/27/2023; EPI\_ISL\_20508078), A(H1N1)pdm09 (A/Wisconsin/67/2022 ; EPI\_ISL\_20508077), and B (A/Austria/1359417/2021; EPI\_ISL\_20508076). Antigenic distinction (or vaccine mismatch) was interpreted per convention based upon ≥8-fold reduction in HI titre against the case virus when compared to the homologous 2025/26 cell-based vaccine reference strain against which ferret antisera were originally raised [41].

We examined age distributions among unvaccinated cases, comparing against unvaccinated controls to standardize for potential sampling differences. In 2025/26, A(H3N2) cases were significantly younger than A(H1N1)pdm09 cases or controls (median age 20 vs. 37 or 37 years) (**Supplementary Figure S1, Table S9**). As context, the median age of SPSN controls has increased since the 2009 A(H1N1)pdm09 pandemic (26 years), notably since 2023/24 (35 years). Whereas the median age of A(H3N2) cases was similar to controls in 2010/11 (33 vs. 31 years), they have been significantly younger starting in 2018/19 (24 vs. 34 years). Influenza A(H1N1)pdm09 cases were significantly younger than controls during the 2009 pandemic (20 vs 26 years), but variously older or more comparable to controls since 2013/14. Influenza A(H3N2) subclade K cases were younger than other non-J.2.4-descendent subclades or controls with more <30-year-olds (median 18 vs. 32 or 37 years; 64% vs. 48% or 38%), whereas A(H1N1)pdm09 subclades D.3.1, D.3.1.1, and controls were comparable (median 34, 37, 37 years; 45%, 40%, 38%) (**Supplementary Table S10**). More influenza B subclade C.3 than C.5 or non-sequenced cases were youth <18 years (median 11 vs. 20 or 33 years; 62% vs. 47% or 34%).

### Vaccine effectiveness

Adjusted VE against medically-attended outpatient ARI is shown by subtype, subclade and age in **Figure 2**. Findings were similar in sensitivity analyses additionally adjusting for sex and comorbidity or excluding COVID-19 infections from controls, generally varying ≤2-4% except with age stratification and reduced sample size (**Supplementary Table S11**).

**Figure 2.**
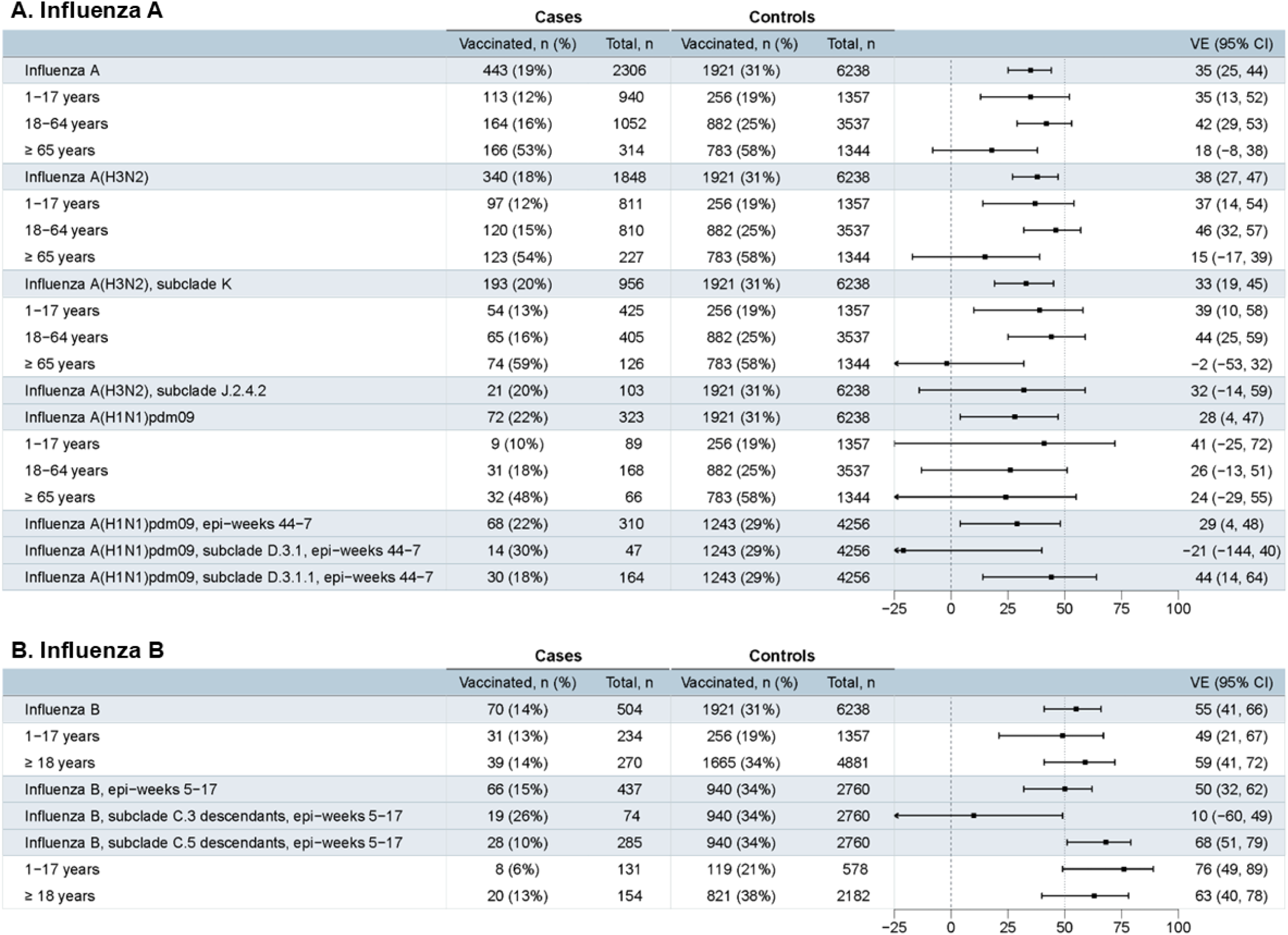
Vaccine effectiveness against influenza, stratified by age, subtype, and subclade, Canadian Sentinel Practitioner Surveillance Network, 26 October 2025 – 02 May 2026 (weeks 44–17). Abbreviations: CI, confidence interval; VE, vaccine effectiveness. Vaccination status per participant/guardian report. Participants vaccinated <2 weeks prior to symptom onset or with unknown vaccination status were excluded. VE was calculated as 1-odds ratio (OR) x 100%. ORs compare percent positivity between vaccinated and unvaccinated participants using logistic regression. Firth’s logistic regression was explored as needed to address small sample size; VE estimates differed by ≤3% (not displayed). Adjusted for age group (1-8, 9-17, 18-49, 50-64, ≥ 65 years), calendar time (bi-weekly epi-week), and province (Alberta, BC, Ontario, Quebec). Additional adjustment for sex and/or comorbidity had minimal impact on VE findings (**Supplementary Table S11**). Subclade K (formerly known as subclade J.2.4.1) and subclade J.2.4.2 viruses share parental subclade J.2.4 clade-defining substitutions T135K (A)(RBS)(−CHO) and K189R (B), plus additional S144N (A)(+CHO), N158D (B), I160K (B) and T328A mutations. Subclade K is differentiated by additional K2N, Q173R (D) and HA2: S49N, while subclade J.2.4.2 is differentiated by F79V. For further details on virological characterization, see **Supplementary Table S1**. Subclade C.3 descendant viruses include subclade C.3.1 (n=65) and C.3.3 (n=9) viruses; there were no detections of parental C.3 viruses or descendant C.3.2 viruses in SPSN specimens. Subclade C.5 descendant viruses include parental subclade C.5 (n=1) and descendant subclades C.5.1 (n=69), C.5.6 (n=114), C.5.6.1 (n=148), and C.5.7 (n=7); there were no detections of subclades C.5.2-C.5.5 in SPSN specimens. For further details on virological characterization, see **Supplementary Table S2.** Overall and subclade-specific VE was assessed for influenza A(H1N1)pdm09 with restriction to epi-weeks 44-07 to capture the period when both subclades co-circulated (see Figure 1). The same applies for restriction of influenza B analyses to epi-weeks 5-17. Displayed cases include two specimens with influenza A(H1N1)pdm09 and A(H3N2) coinfection, and one specimen with influenza A(H3N2) and influenza B coinfection.

VE against A(H3N2) was 38% (95% confidence interval: 27%–47%), similar against predominant subclade K (33%) and subclade J.2.4.2 (32%) (**Figure 2**). A(H3N2) and subclade K VE were lowest in adults ≥65 years at 15% (−17%–39%) and −2% (−53%–32%), respectively.

VE against A(H1N1)pdm09 was 28% (4%–47%), higher against predominant subclade D.3.1.1 (44%; 14%–64%) than parental subclade D.3.1 (−21%; −144%–40%), but with wide confidence intervals (**Figure 2**). With age stratification, VE was highest in ≤17-year-olds at 41% (−25%–72%).

VE against influenza B was 55% (41%–66%), higher against subclade C.5 at 68% (51%–79%) than C.3 at 10% (−60%–49%) (**Figure 2**). VE was 49% (21%–67%) in youth ≤17 years and 59% (41%–72%) in adults ≥18 years.

We explored repeat vaccination effects in **Supplementary Table S12**. With updated A(H3N2) vaccine in 2025/26, VE was similar for current season’s only (41%) and current plus prior (37%) seasons’ vaccinees. With identical A(H1N1)pdm09 vaccine since 2023/24, VE was higher for current season’s only than current plus prior seasons’ vaccinees (59% vs. 36%, respectively). With identical influenza B/Victoria vaccine since 2022/23, VE was also higher for current-season-only than current-plus-prior-season vaccinees (62% vs. 52%, respectively).

## Discussion

In this end-of-season analysis the Canadian SPSN estimates that, relative to unvaccinated individuals, the 2025/26 influenza vaccine reduced the risk of medically-attended outpatient illness due to A(H3N2) by more than one-third, including against predominant vaccine-mismatched subclade K; A(H1N1)pdm09 by more than one-quarter against vaccine-matched subclades D.3.1 and D.3.1.1, with paradoxical variation between subclades; and due to B/Victoria by more than half, with variation between vaccine-mismatched subclade C.3 and vaccine-matched subclade C.5. Overall, the 2025/26 season highlights inconsistency between conventional laboratory indicators of vaccine match or mismatch and epidemiological observations of vaccine performance.

Emergence of subclade K generated substantial pre-season concern. With antigenic site A mutations affecting viral glycosylation and site B mutations affecting pivotal positions 158 and 189, subclade K was widely considered an important drift variant. As further reinforced among SPSN case viruses, antigenic characterization showed ≥64-fold difference in HI titres against subclade K relative to the egg-based vaccine strain, far exceeding the conventional ≥8-fold threshold defining vaccine-mismatch [1,9,19]. Antigenic characterization using first-infection ferret antisera, however, does not reflect the complex immunological interactions underpinning vaccine response in humans, including potent childhood imprinting, accumulated lifetime back-boosting exposures, and varying profiles of pre-existing immunity [20]. As described previously, glycosylation changes in circulating A(H3N2) viruses in the 1990s likely resulted in immunodominance shift from antigenic site A (pre-1997 birth cohorts) to antigenic site B (post-1997 birth cohorts) [1,7]. Loss of the N122 glycan among subclade J.2 and descendant viruses, including the 2025/26 vaccine strain and subclade K, may have exposed a previously glycan-shielded site A epitope benefiting older cohorts with site A immuno-dominant orientation [7,21]. With recently-acquired site B mutations, subclade K may have posed greater risk to site B imprinted and oriented youth, as suggested by age distributions among unvaccinated SPSN participants [1,7]. The extent to which virological evolution contributes to an overall younger A(H3N2) (but not A(H1N1)pdm09) profile relative to controls in this outpatient setting warrants more granular investigation, recognizing median age is a crude indicator and SPSN controls may be older than historic, notably since the COVID-19 pandemic. Unlike immunologically-naïve ferrets, human adults show cross-reactive antibody against subclade K, further boosted by 2025/26 vaccination [22–25]. Also unlike ferrets, human adult antibody responses may not be so focused toward site B, but may encompass more accumulated and broadly compensatory recognition of sites C, D, or E less affected by subclade K mutations [26].

Our 2025/26 A(H3N2) VE was higher than expected given antigenic characterization of vaccine mismatch. The end-of-season VE against subclade K (33%; similar to mid-season 37% [7]) is within range of prior SPSN A(H3N2) VE estimates of the past two decades, spanning 30-60% in 12/16 seasons [12], including 2024/25 (40%) and 2023/24 (32%) when most viruses in Canada were vaccine-matched antigenically (89% and 94%, respectively [3,27]). Despite adjuvanted or high-dose influenza vaccines for older adults, 2025/26 A(H3N2) VE was lower among ≥65-year-olds (15%) than 18-64-year-olds (46%), although age-related patterns varied elsewhere in mid-season outpatient estimation [8,9,28,29]. Point estimates spanned 10% (Denmark) to 37% (United States [US]) among older adults and more widely from 12% (US) to 61% (Denmark) among younger adults [8,9,28,29], potentially reflecting site-specific differences (e.g., available vaccines). Our antigenic characterization demonstrated greater mismatch between subclade K and egg-based vaccine, the latter including a D186A (B) egg-adaptation mutation not found in cell-based vaccine. Human immunogenicity studies suggest lower subclade K cross-reactivity following egg-based versus recombinant- [25] and potentially cell-based vaccine administration [24]. The egg-based 2025/26 live-attenuated influenza vaccine (LAIV) is a J.2.2 strain distinguished from inactivated J.2 vaccine (and subclade K) by S124N (A). LAIV lacks D186A (B) but has other egg-adaptation mutations (S219Y (D) and I226M (D)(RBS)). Ecologically, 2025/26 A(H3N2) VE in children was higher where there was greater LAIV use. With <1% of available product being LAIV, A(H3N2) VE in children was lower in our network (37%) and in Beijing (20%) compared to primary care networks across Europe (VE: 53%; LAIV: >30% in children) and the United Kingdom (UK)(VE: 61%; LAIV: >90% in children)[8,29]. In California, Zhu et al. also report higher 2025/26 VE against any influenza using LAIV (55%) vs inactivated vaccine (39%) during an A(H3N2)-predominant period, albeit without direct typing/subtyping of case viruses [30].

Our 2025/26 A(H1N1)pdm09 VE was lower than expected given antigenic characterization of vaccine match. In fact, our A(H1N1)pdm09 VE of 28% is the lowest of any prior end-of-season A(H1N1)pdm09 VE reported by the SPSN since 2006/07, ranging seasonally from 37% (2024/25) to 92% (2006/07) and generally ≥50% (11/14 seasons)[12]. The 2025/26 A(H3N2) VE exceeded A(H1N1)pdm09 VE which is itself historically exceptional among SPSN estimates [10,12]. To our knowledge, only one other study has reported 2025/26 A(H1N1)pdm09 VE [8]. Estimates through mid-January from primary care networks in the UK, broader Europe, and Denmark (32%, 34%, and 35%, respectively) were similar to ours [8]. Among SPSN participants, almost all A(H1N1)pdm09 viruses were clade 5a.2a.1, with a mix of subclades D.3.1 and descendant D.3.1.1 earlier in the season followed by later D.3.1.1 predominance. Antigenically-characterized viruses were almost entirely matched to clade 5a.2a.1 vaccine, similar for both circulating subclades and in relation to cell- (subclade C.1.1) or egg-based (subclade D) vaccine strains. Nonetheless, we observed subclade-specific VE differences, unexpectedly lower against the parental D.3.1 (nil) than descendant D.3.1.1 (44%) subclade.

Discordant antigenicity and VE findings may be explained by ferret antisera preferentially recognizing immunodominant antigenic site Sa, whereas recent A(H1N1)pdm09 viruses have accumulated mutations across other antigenic sites potentially more relevant to human immunity and less detectable by ferret-based assays [31]. Human serology studies for currently circulating A(H1N1)pdm09 strains remain limited, but findings presented at the September 2025 WHO vaccine composition meeting showed progressively (and perhaps paradoxically) improved human antibody reactivity relative to D.3.1 following acquisition of additional D.3.1.1-defining substitutions R113K and A139D (Ca2) + E283K [32], consistent with our higher subclade-specific VE against D.3.1.1. We previously observed paradoxically lower VE against A(H1N1)pdm09 vaccine clade 5a.2a.1 compared to clade 5a.2a viruses in 2023/24 [14,33,34], with potential mechanisms including the R142K (Ca2) reversion in the egg-adapted 5a.2a.1 vaccine strain, but not circulating 5a.2a.1 viruses, and/or mutation at position 216 of both vaccine and circulating 5a.2a.1 viruses potentially disrupting glycosylation patterns and imprint-related effects [14,35]. With unchanged vaccine since 2023/24, negative interference associated with repeat vaccination may also contribute as indicated by lower 2025/26 A(H1N1)pdm09 VE estimates among current-plus-prior (36%) versus current-season-only (59%) vaccinees.

Our 2025/26 influenza B VE (55%) was also amongst the lowest of historic SPSN B/Victoria estimates since 2010/11, ranging 51% (2010/11) to 78% (2012/13) and exceeding 60% in most (5/7) prior seasons [12]. We genetically characterized more than three-quarters of our contributing influenza B case viruses, with subclade C.5 descendants predominating overall and lesser subclade C.3 co-circulation. Compared to cell-based vaccine, all SPSN subclade C.5 viruses were vaccine-matched but all subclade C.3 were vaccine-mismatched, consistent also with human serological data [36,37]. Although antigenic characterization using egg-based vaccine reference virus and homologous ferret antisera suggested vaccine mismatch to both subclades, using egg-based antisera heterologous to cell-based vaccine reference, more akin to cell-propagated SPSN test viruses, restored conclusions of egg-based vaccine match to C.5 but not C.3. Unlike subclade C.5 and descendant viruses, C.3 descendants harbor the D197N (190-helix)(RBS)(+CHO) mutation conferring gain-of-glycosylation, with subclade C.3 activity driven by descendant subclade C.3.1, a reassortment between C.3 (HA, PB1, PB2, M segments) and C.5.1 (NA, NP, PA, NS segments) [36]. In a cohort of 50 adults, significant decrease in pre- and post-vaccination neutralizing antibody titres, and fold-rises, were observed against C.3.1 relative to C.3 and C.5.1 [36]. Responses against C.3.1 viruses were rescued with N197D reversion, supporting a major role for the N197 glycan in antigenic distinction [36]. We provide further epidemiological support for these virological observations, showing lower VE against (N197) subclade C.3 descendants (10%) than (E197) C.5 descendants (68%). Given unchanged B/Victoria vaccine since 2022/23, lower VE may also align with repeat vaccination effects. Although the influenza B/Victoria component has been updated to subclade C.3.1 for the NH 2026/27 vaccine, a T199I mutation in the egg-based strain confers loss of the N197 glycan present in C.3 descendant viruses, suggesting vaccine mismatch may be an ongoing issue should they continue to circulate.

Analyses from the US VISION network, spanning to epi-week 10, generally align with our age-stratified VE findings against influenza B in participants ≥18 years (VISION: 63%; SPSN: 59%) and ≤17 years (VISION: 38%; SPSN: 49%) [38]. In studies reporting greater contribution of subclade C.3 descendants,including ∼85% in the outpatient US Influenza VE Network (per direct sequencing of contributing case viruses [28]) and a hospital network in Japan (per surveillance data [39]), reported VE was even lower among both children ≤17 years (20%, 25%, respectively) and adults ≥18 years (23% in the US). As in recent seasons, approximately half of SPSN influenza B cases in 2025/26 were children, including among unvaccinated participants. Stratifying by sequencing result showed non-sequenced vs. sequenced influenza B case viruses were generally older, a pattern not replicated among influenza A cases. Non-sequenced specimens include those for whom sequencing failed or was not attempted due to low viral load. We postulate the non-sequenced subset may have been older on account of pre-existing immunity to influenza B, suppressing viral load. Subclade C.3 were younger than C.5 cases (median 11 vs. 20 years), consistent with patterns shown in a subset of influenza B cases from Johns Hopkins Hospital System in 2024/25, suggested by Akin et al. (2026) to reflect more concentrated impact of antigenically drifted C.3.1 among younger more immunologically-naïve children [36].

Like other observational methods to estimate VE, our study has limitations including potential for residual bias and confounding. Limited sample size in sub-stratified analyses, especially by subclade, age and prior vaccination status, limits statistical power and introduces instability and uncertainty warranting caution. We cannot explore differences by vaccine type with ≥90% of influenza vaccines in SPSN provinces being egg-based and inactivated. Generalization of findings to other settings with a different mix of vaccines, circulating variants or other relevant context (e.g., inpatient/outpatient, age, exposure or outcome risks) also requires caution.

## Conclusion

In this end-of-season analysis, the Canadian SPSN estimates the 2025/26 influenza vaccine reduced the risk of medically-attended ARI by over a third for influenza A(H3N2), a quarter for influenza A(H1N1)pdm09, and half for influenza B. The 2025/26 season reinforces the importance of integrated genetic, antigenic and epidemiologic VE monitoring, including subclade-specific. Conventional antigenic characterization of vaccine match or mismatch based on ferret antisera is insufficient, requiring incorporation of other indicators (e.g., human serology, immunogenicity) and innovations (e.g., deep mutational scanning, high-throughput sequencing-based neutralization assays) to monitor emergence and gauge importance of immune escape variants. Ultimately, in vitro markers may be helpful intermediaries to track viral evolution and anticipate impact but their linkage to epidemiological estimates of subclade-specific VE is critical for confirmation and complete understanding.

## Conflicts of interest

DMS is Principal Investigator on grants received to her institution from the Public Health Agency of Canada, Canadian Institutes of Health Research, and the Pacific Public Health Foundation. JAD reports funding from Alberta Health paid to his institution. ANJ reports funding from the Canadian Institutes of Health Research, Genome Canada, and Genome British Columbia paid to her institution, and financial support from the Canadian Institutes of Health Research and Canadian Association for Clinical Microbiology and Infectious Diseases for travel expenses, with none of the above pertaining to the current study. ANJ also reports committee membership in the Canadian Association for Clinical Microbiology and Infectious Diseases (no payments made) and the British Columbia Diagnostic Accreditation Program (payments made to ANJ). SC reports funding from the Public Health Agency of Canada paid to her institution, but not pertaining to the current study. Other authors have no conflicts of interest to declare.

## Funding

This work was supported by funding from the BC Ministry of Health, Pacific Public Health Foundation, Alberta Health and Wellness, Public Health Ontario, the Ministère de la santé et des services sociaux du Québec, the Canadian Institutes of Health Research [grant number FP5-203220], and the Public Health Agency of Canada [grant arrangement number 2324-HQ-000038]. The views expressed herein do not necessarily represent the view of the Public Health Agency of Canada. Funders had no role in data analysis, interpretation or the decision to publish.

## Ethics

Ethical approval was obtained or waived in the four participating provinces as follows. In Alberta, the Conjoint Health Research Ethics Board of the University of Calgary gave ethical approval for this work (REB150587). In British Columbia, both the Clinical and Behavioural Research Ethics Boards of the University of British Columbia waived ethical approval for this work because such evaluations are considered within the core public health mandate of the BC Centre for Disease Control (BCCDC). In Ontario, the Ethics Review Board of Public Health Ontario determined this work did not require ongoing review and waived ethical approval for this work, as the activities are considered routine public health practice in fulfilment of Public Health Ontario’s legislated mandate, and not research. In Québec, the Research Ethics Board of the Centre hospitalier universitaire de Québec waived ethical approval for this work as such evaluations are similarly considered part of core public health surveillance.

## Acknowledgements

The authors gratefully acknowledge the contribution of sentinel sites whose regular submission of specimens and data provide the basis of our analyses. We wish to acknowledge the administrative, coordination, data entry and/or management support in participating provinces including: Gabriel Canizares for provincial and national coordination at the British Columbia Centre for Disease Control; Emilie Toews, Tolulope Ogunniyi, and Anamika Kambo for TARRANT in Alberta; Kara Gill and Kanti Pabbaraju for coordinating sequencing at the Alberta Provincial Laboratory for Public Health; Julie Carange and Lyne Désautels for Institut national de santé publique du Québec, Josiane Rivard and Stéphanie Grenier for Centre de recherche du CHU de Québec-Université Laval; and Mandy Kwok and the genomics and bioinformatics team for Public Health Ontario. We wish to thank those who provided additional laboratory and technical support at the British Columbia Centre for Disease Control Public Health Laboratory, the Alberta Provincial Laboratory for Public Health, Public Health Ontario Laboratory, the Laboratoire de santé publique du Québec, and the National Microbiology Laboratory. We thank the WHO Collaborating Centre for Reference and Research on Influenza in the UK (The Francis Crick Institute) for providing us with the egg-based and cell-based vaccine viruses and the homologous egg-based vaccine virus ferret antisera used in our HI assays. We also gratefully acknowledge the authors, originating laboratories, and submitting laboratories of the vaccine reference virus strains for sharing those via the GISAID Initiative, which we rely on for our reference sequences.

## Data availability statement

Sequencing data for included viruses that met provincial/national criteria for upload and their submitting/contributing laboratories can be found on GISAID using the Epi Set ID: EPI_SET_260819zv (https://doi.org/10.55876/gis8.260819zv).

## Notes

### Author Declarations

Ethical approval was obtained or waived in the four participating provinces as follows. In Alberta, the Conjoint Health Research Ethics Board of the University of Calgary gave ethical approval for this work (REB150587). In British Columbia, both the Clinical and Behavioural Research Ethics Boards of the University of British Columbia waived ethical approval for this work because such evaluations are considered within the core public health mandate of the BC Centre for Disease Control (BCCDC). In Ontario, the Ethics Review Board of Public Health Ontario determined this work did not require ongoing review and waived ethical approval for this work, as the activities are considered routine public health practice in fulfilment of Public Health Ontario's legislated mandate, and not research. In Quebec, the Research Ethics Board of the Centre hospitalier universitaire de Quebec waived ethical approval for this work as such evaluations are similarly considered part of core public health surveillance.

