## Supplementary materials for "2025/26 subclade-specific influenza A and B vaccine effectiveness estimates and discordant laboratory-based indicators of vaccine match or mismatch"

**Supplementary Methods. Antigenic characterization of viruses included in vaccine effectiveness analyses**

Hemagglutination inhibitions (HI) assays were conducted at the National Microbiology Laboratory (NML) using cell-based and egg-based vaccine reference strains provided by the Francis Crick Institute. All contributing case (test) viruses of the Sentinel Practitioner Surveillance Network (SPSN) were cultured in Madin-Darby canine kidney (MDCK) cells, with fold reductions variously assessed in relation to vaccine reference titres measured as detailed below. In all scenarios, regardless of vaccine reference virus used to raise ferret antisera or as comparator strain, antigenic distinction was defined as $\geq$8 fold-reduction in HI titre against the cell-propagated SPSN case (test) virus, relative to titres generated against the comparator vaccine reference virus [1]. Varying assay approaches included:

1. Primary homologous characterization (per convention): ferret antisera were raised against cell-based vaccine viruses (at the NML), which were then used to measure homologous vaccine reference titers (i.e., cell-raised ferret antisera and cell-based vaccine virus) against which fold-reductions in titres against the cell-propagated SPSN test viruses were assessed.
2. Secondary homologous characterization (per convention): as also commonly undertaken, ferret antisera were raised against egg-based vaccine viruses (at the Francis Crick Institute), which were then used per convention to measure homologous vaccine reference titers (i.e., egg-raised ferret antisera and egg-based vaccine virus) [2], against which fold-reductions in titres against the cell-propagated SPSN test viruses were also assessed in sensitivity analyses.
3. Sensitivity heterologous characterization (exploratory): to better replicate the egg-based vaccine but cell-propagated SPSN case (test) virus scenario, ferret antisera were raised against egg-based vaccine viruses (at the Francis Crick Institute) which were then used to explore heterologous vaccine reference titres (i.e., egg-raised ferret antisera and cell-based vaccine virus) against which fold-reductions in titres against the cell-propagated SPSN test viruses were also assessed in sensitivity analyses.

| Viruses used to generate comparator reference titres for HI assays (from Francis Crick Institute) | Viruses used to raise ferret antisera | |
|---|---|---|
| | Cell-based vaccine virus (NML generated antisera) | Egg-based vaccine virus (Francis Crick Institute generated antisera) |
| Cell-based vaccine virus | 1. Primary homologous (per convention) | 3. Sensitivity heterologous (exploratory) |
| Egg-based vaccine virus | —[a] | 2. Secondary homologous (per convention) |

[a] Not undertaken because not relevant to the purpose of assessing in relation to cell-based SPSN case (test) viruses

**Supplementary Table S1.** Genetic distribution of influenza A case viruses (n=2169) included in vaccine effective analyses, Canadian Practitioner Surveillance network, 26 October 2025 – 02 May 2026 (week 44-17).

| **Genetic clade, as defined by ECDC** [3] **based upon specific HA amino acid substitutions**<br>+ additional substitutions by subclade or uniquely identified (antigenic site) | **NextStrain subclade** [5,6] | **BC** | **Alberta** | **Ontario** | **Québec** | **TOTAL** |
|---|---|---|---|---|---|---|
| **Influenza A(H3N2), N (case viruses)**[a]<br>Case viruses successfully sequenced, n (% n/N) | | **225**<br>**175 (78%)** | **247**<br>**178 (72%)** | **1069**<br>**570 (53%)** | **307**<br>**176 (57%)** | **1848**[b]<br>**1099 (59%)** |
| **2a.3a.1 = 2a.3a + I140K (A) + I223V** | **J** | **21 (12%)** | **12 (7%)** | **90 (16%)** | **21 (12%)** | **143 (13%)** |
| + N122D (A)(-CHO) + K276E (C) | J.2[c] | | | | 6 | 6 |
| + S124N (A) | J.2.2 | 1 | | 2 | | 3 |
| + E83D (E) + I214T (D) + HA2: V18M | | 2 | 9 | | | 11 |
| + N158K (B) + K189R (B) + HA2: S49N | J.2.3 | 1 | 1 | | | 2 |
| + S54N (C) + G78S (E) + S145N (A) + N216H (D) | | 2 | 1 | 1 | 12 | 16 |
| + T135K (A)(RBS)(-CHO) + K189R (B) | J.2.4 | | | 1 | 1 | 2 |
| + F79V + S144N (A)(+CHO) + N158D (B) + I160K (B) + T328A | J.2.4.2 | 15 | 1 | 85 | 2 | 103 |
| **J.2.4 + K2N + S144N (A)(+CHO) + N158D (B) + I160K (B) + Q173R (D) + T328A + HA2: S49N** | **K**<br>(formerly J.2.4.1) | **154 (88%)** | **166 (93%)** | **481 (84%)** | **155 (88%)** | **956 (87%)** |
| + N144D (A)(-CHO) + K278E (C) | | 3 | 30 | | | 33 |
| + S145N (A)[d] | | 5 | | 7 | 1 | 13 |
| + S198P (B) | | | | 4 | 10 | 14 |
| **Influenza A(H1N1)pdm09, N (case viruses)**[a]<br>Case viruses successfully sequenced, n (% n/N) | | **54**<br>**48 (895)** | **45**<br>**44 (98%)** | **132**<br>**60 (46%)** | **92**<br>**70 (76%)** | **323**[e]<br>**222 (69%)** |
| **5a.2** = 5a + K130N + N156K (Sa) + L161I (Sa) + V250A + HA2: E179D | C | | | | | |
| **5a.2a = 5a.2 + K54Q + A186T (Sb) + Q189E (Sb) + E224A (RBS) + E259K + K308R** | **C.1** | | | **1 (2%)** | **3 (4%)** | **4 (2%)** |
| + S83P + T120A + K169Q (Ca1) + HA2: I91V + I183T | C.1.9.3 | | | 1 | 3 | 4 |
| **5a.2a.1 = 5a.2a + P137S (Ca2) + K142R (Ca2) + D260E + T277A + HA2: E29D + I91V + N124H** | **C.1.1**[c] | **48 (100%)** | **44 (100%)** | **59 (98%)** | **67 (96%)** | **218 (98%)** |
| + T216A | D[c] | | | | | |
| + T120A + HA2: I45V + I133T + V193A | D.3.1 | 24 | 8 | 7 | 8 | 47 |
| + R113K + A139D (Ca2) + E283K + K302E | D.3.1.1 | 18 | 19 | 39 | 54 | 130 |
| + D139N (Ca2)[f] | | 4 | 14 | 4 | 3 | 25 |
| + R205K (Ca1) | | 2 | 3 | 9 | 2 | 16 |

A(H3N2) colour coding aligns with Figure 1 of the main manuscript. Additional substitutions included if located in antigenic or receptor binding sites or involved in gain or loss of glycosylation AND present in >1% of A(H3N2) sequenced viruses or >2% of A(H1N1)pdm09 sequenced viruses.

---

[a] Influenza A(H3N2) substitutions are relative to A/Massachusetts/18/2022 (EPI_ISL_16998756) 2a.3a.1 (subclade J) reference virus and A(H1N1)pdm09 substitutions are relative to A/Wisconsin/588/2019 (EPI_ISL_404460) 5a.2 (subclade C) reference virus [4]

[b] Includes one virus with influenza A(H3N2) + influenza B co-infection counted twice, but only successfully sequenced as B/Victoria subclade C.5.6. Also includes two viruses with influenza A(H3N2) + influenza A(H1N1)pdm09 co-infection, both counted twice. One specimen sequenced as A(H1N1)pdm09 subclade D.3.1 and the other sequenced as A(H3N2) subclade K.

[c] The 2025/26 influenza A(H3N2) vaccine strain was subclade J.2 (cell- and egg-based); influenza A(H1N1)pdm09 vaccine strains were subclade C.1.1 (cell-based) and subclade D (egg-based).

[d] Viruses do not cluster together phylogenetically

[e] Includes two viruses with influenza A(H3N2) + influenza A(H1N1)pdm09 co-infection, both counted twice. One specimen sequenced as A(H1N1)pdm09 subclade D.3.1 and the other sequenced as A(H3N2) subclade K.

[f] One sample acquired D139N mutation separately from cluster

**Supplementary Table S2.** Genetic distribution of influenza B case viruses (n=504) included in vaccine effective analyses, Canadian Practitioner Surveillance network, 26 October 2025 – 02 May 2026 (week 44-17).

| **Genetic clade, as defined by ECDC** [3] **based upon specific HA amino acid substitutions**<br>+ additional substitutions by subclade or uniquely identified (antigenic site) | **NextStrain subclade** [5,6] | **BC** | **Alberta** | **Ontario** | **Québec** | **TOTAL** |
|---|---|---|---|---|---|---|
| **Influenza B/Victoria, N (case viruses)**[a]<br>Case viruses successfully sequenced, n (% n/N) | | **75**<br>**65 (87%)** | **128**<br>**115 (90%)** | **226**<br>**169 (75%)** | **75**<br>**64 (85%)** | **504**[b]<br>**413 (82%)** |
| **V1A.3a.2** = V1A.3a + A127T (120-loop) + P144L (150-loop) + K203R (190-helix)(RBS) | C[c] | | | | | |
| **+ E128K (120-loop) + A154E + S208P** | **C.3** | **15 (23%)** | **3 (3%)** | **49 (29%)** | **7 (11%)** | **74 (18%)** |
| + D197N (190-helix)(RBS)(+CHO) + P208S (reversion) | C.3.1 | 10 | 3 | 41 | 6 | 60 |
| + N123K (120-loop) | | | | 5 | | 5 |
| + D197N (190-helix)(RBS)(+CHO) + S255P + I267V | C.3.3 | 5 | | 3 | 1 | 9 |
| **+ D197E (190-helix)(RBS)** | **C.5** | **50 (77%)** | **112 (97%)** | **120 (71%)** | **57 (89%)** | **339 (82%)** |
| + E183K | C.5.1 | | | | | |
| + E128K (120-loop) | | | | 7 | 4 | 11 |
| + H122Q (120-loop) | | 5 | 35 | | 1 | 41 |
| + N126S (120-loop) + A202V (190-helix)(RBS) + HA2: R152K | | 11 | 3 | 3 | | 17[d] |
| + D129N (120-loop) | C.5.6 | 3 | 10 | 70 | 31 | 114 |
| + T37I + E128D (120-loop) + T199A (190-helix)(RBS) | C.5.6.1 | 3 | 1 | 4 | 10 | 18 |
| + P241Q (230-region)(RBS) | | 20 | 51 | 30 | 8 | 109 |
| + N171D + E198K (190-helix)(RBS) | | 4 | 12 | 4 | 1 | 21 |
| + E128G (120-loop) + E183K | C.5.7 | 4 | | 1 | 2 | 7 |

B/Victoria colour coding aligns with Figure 1 of the main manuscript. Additional substitutions included if located in antigenic or receptor binding sites or involved in gain or loss of glycosylation AND present in >1% of sequenced viruses.

[a] Influenza B substitutions are relative to A/Austria/1359417/2021 (EPI_ISL_983345) V1A.3a.2 (subclade C) reference virus [4]

[b] Includes one virus with influenza A(H3N2) + influenza B co-infection; counted twice. Specimen sequenced as B/Victoria subclade C.5.6

[c] The 2025/26 influenza B vaccine strain was subclade C (cell- and egg-based).

[d] One sample with K128E reversion

**Supplementary Table S3.** Antigenic characterization of a subset of influenza A(H3N2) viruses included in vaccine effectiveness analysis that were tested by hemagglutination inhibition (HI) assay using various combinations of egg- and cell-derived virus and antisera, Canadian Practitioner Surveillance Network (SPSN), 26 October 2025 – 2 May 2026 (epi-weeks 44-17) (n=104).

| Subclade | Reference vaccine virus[a] used to measure comparator titre | Reference vaccine virus used to generate ferret antisera | Total tested (N) | Antigenic result (n) | | Fold-difference among antigenically distinct (n) | | | |
|---|---|---|---|---|---|---|---|---|---|
| | | | | Similar | Distinct | 8 | 16 | 32 | ≥64 |
| J.2 | **Cell (homologous)** | **Cell** | 4 | **4** | **-** | | | | |
| | Egg (homologous) | Egg | | 4 | - | | | | |
| | Cell (heterologous) | Egg | | 4 | - | | | | |
| J.2.2 | **Cell (homologous)** | **Cell** | 4 | **4** | **-** | | | | |
| | Egg (homologous) | Egg | | 4 | - | | | | |
| | Cell (heterologous) | Egg | | 4 | - | | | | |
| J.2.3 | **Cell (homologous)** | **Cell** | 4 | **-** | **4** | **-** | **-** | **-** | **4** |
| | Egg (homologous) | Egg | | - | 4 | - | - | - | 4 |
| | Cell (heterologous) | Egg | | - | 4 | - | - | - | 4 |
| J.2.4 | **Cell (homologous)** | **Cell** | 2 | **1** | **1** | **1** | **-** | **-** | **-** |
| | Egg (homologous) | Egg | | - | 2 | 1 | 1 | - | - |
| | Cell (heterologous) | Egg | | 1 | 1 | 1 | - | - | - |
| J.2.4.2 | **Cell (homologous)** | **Cell** | 10 | **-** | **10** | **-** | **6** | **4** | **-** |
| | Egg (homologous) | Egg | | - | 10 | - | - | 1 | 9 |
| | Cell (heterologous) | Egg | | - | 10 | - | 1 | 1 | 8 |
| K[b] | **Cell (homologous)** | **Cell** | 80 | **-** | **80** | **19** | **52** | **9** | **-** |
| | Egg (homologous) | Egg | | - | 80 | - | - | 1 | 79 |
| | Cell (heterologous) | Egg | | - | 80 | - | 1 | 38 | 41 |

All SPSN case (test) viruses were cell-propagated in each HI assay approach as detailed in **Supplementary Methods**. Bolding indicates conditions as per primary antigenic characterization methods but here restricted to the subset assessed by all three HI assay approaches.

[a] Cell-based vaccine strain was A/District of Columbia/27/2023 (EPI_ISL_20508078) and egg-based vaccine strain was A/Croatia/10136RV/2023 (EPI_ISL_20508079). Both were clade 2a.3a.1, subclade J.2 with additional S145N (A) substitution, and the egg-based vaccine strain acquiring a D186A (B) mutation relative to the cell-based vaccine strain.
[b] Formerly known as subclade J.2.4.1.

**Supplementary Table S4.** Antigenic characterization of a subset of influenza A(H1N1)pdm09 viruses included in vaccine effectiveness analysis that were tested by hemagglutination inhibition assay using various combinations of egg- and cell-derived virus and antisera, Canadian Practitioner Surveillance Network, 26 October 2025 – 02 May 2026 (epi-weeks 44-17) (n=67).

| Subclade | Reference vaccine virus[a] used to measure comparator titre | Reference vaccine virus used to generate ferret antisera | Total tested (N) | Antigenic result (n) | | Fold-difference among antigenically distinct (n) | | | |
|---|---|---|---|---|---|---|---|---|---|
| | | | | Similar | Distinct | 8 | 16 | 32 | ≥64 |
| D.3.1 | **Cell (homologous)** | **Cell** | 27 | **26** | **1** | **1** | **-** | **-** | **-** |
| | Egg (homologous) | Egg | | 27 | - | | | | |
| | Cell (heterologous) | Egg | | 27 | - | | | | |
| D.3.1.1 | **Cell (homologous)** | **Cell** | 38 | **38** | **-** | | | | |
| | Egg (homologous) | Egg | | 37 | 1 | 1 | - | - | - |
| | Cell (heterologous) | Egg | | 37 | 1 | - | 1 | - | - |
| unknown | **Cell (homologous)** | **Cell** | 2 | **2** | **-** | | | | |
| | Egg (homologous) | Egg | | 2 | - | | | | |
| | Cell (heterologous) | Egg | | 2 | - | | | | |

All SPSN case (test) viruses were cell-propagated in each HI assay approach as detailed in **Supplementary Methods**. Bolding indicates conditions as per primary antigenic characterization methods but here restricted to the subset assessed by all three HI assay approaches.

[a] Cell-based vaccine strain was A/Wisconsin/67/2022 (EPI_ISL_20508077) belonging to clade 5a.2a.1, subclade C.1.1, and egg-based vaccine strain was A/Victoria/4897/2022 (EPI_ISL_20508080) belonging to clade 5a.2a.1, subclade D with additional T216A mutation. Additionally, the egg-based strain acquired a Q223R (RBS) egg-adaptation mutation and an R142K (Ca2) high-growth reassortant adaptation (reversion of a clade 5a.2a.1 defining mutation).

**Supplementary Table S5.** Antigenic characterization of a subset of influenza B viruses included in vaccine effectiveness analysis that were tested by hemagglutination inhibition assay using various combinations of egg- and cell-derived virus and antisera, Canadian Practitioner Surveillance Network, 26 October 2025 – 02 May 2026 (epi-weeks 44-17) (n=93).

| Subclade | Reference vaccine virus[a] used to measure comparator titre | Reference vaccine virus used to generate ferret antisera | Total tested (N) | Antigenic result (n) | | Fold-difference among antigenically distinct (n) | | | |
|---|---|---|---|---|---|---|---|---|---|
| | | | | Similar | Distinct | 8 | 16 | 32 | ≥64 |
| C.3.1 | **Cell (homologous)** | **Cell** | 10 | **-** | **10** | **-** | **8** | **2** | **-** |
| | Egg (homologous) | Egg | | - | 10 | - | - | - | 10[b] |
| | Cell (heterologous) | Egg | | - | 10 | 10[c] | - | - | - |
| C.3.3 | **Cell (homologous)** | **Cell** | 5 | **-** | **5** | **1** | **4** | **-** | **-** |
| | Egg (homologous) | Egg | | - | 5 | - | - | - | 5[b] |
| | Cell (heterologous) | Egg | | - | 5 | 5[c] | - | - | - |
| C.5.1 | **Cell (homologous)** | **Cell** | 17 | **17** | **-** | | | | |
| | Egg (homologous) | Egg | | - | 17 | - | 4 | 13 | - |
| | Cell (heterologous) | Egg | | 17 | - | | | | |
| C.5.6 | **Cell (homologous)** | **Cell** | 31 | **31** | **-** | | | | |
| | Egg (homologous) | Egg | | - | 31 | - | 30 | 1 | - |
| | Cell (heterologous) | Egg | | 31 | - | | | | |
| C.5.6.1 | **Cell (homologous)** | **Cell** | 22 | **22** | **-** | | | | |
| | Egg (homologous) | Egg | | - | 22 | - | 12 | 10 | - |
| | Cell (heterologous) | Egg | | 22 | - | | | | |
| C.5.7 | **Cell (homologous)** | **Cell** | 4 | **4** | **-** | | | | |
| | Egg (homologous) | Egg | | - | 4 | - | 4 | - | - |
| | Cell (heterologous) | Egg | | 4 | - | | | | |
| unknown | **Cell (homologous)** | **Cell** | 4 | **4** | **-** | | | | |
| | Egg (homologous) | Egg | | - | 4 | - | 1 | 3 | - |
| | Cell (heterologous) | Egg | | 4 | - | | | | |

All SPSN case (test) viruses were cell-propagated in each HI assay approach as detailed in **Supplementary Methods**. Bolding indicates conditions as per primary antigenic characterization methods but here restricted to the subset assessed by all three HI assay approaches.

[a] Cell-based (EPI_ISL_20508076) and egg-based (EPI_ISL_20508081) vaccine strains were both A/Austria/1359417/2021 belonging to clade V1A.3a.2, subclade C. Both cell- and egg-based viruses acquired an additional G141R mutation.

[b] All specimens had fold-differences ≥128 against egg-based vaccine and antisera

[c] Analysis only permitted fold-changes to be derived as 8, or >8; no further distinction was possible

**Supplementary Table S6.** Participant profile, influenza A(H1N1)pdm09 analyses, Canadian Sentinel Practitioner Surveillance Network, 26 October 2025 – 02 May 2026 (epi-weeks 44–17).

| Characteristics | All ARI participants (column %) | | | | | | Influenza vaccinated[a] (row %) | | | | | |
|---|---|---|---|---|---|---|---|---|---|---|---|---|
| | Overall | | Influenza A(H1N1)pdm09 cases | | Influenza controls | | Overall | | Influenza A(H1N1)pdm09 cases | | Influenza controls | |
| | n | % | n | % | n | % | n | % | n | % | n | % |
| N (row %) | 6561 | 100 | 323 | 5 | 6238 | 95 | 1993 | 30 | 72 | 22 | 1921 | 31 |
| **Age group (years)** [b] | | | | | | | | | | | | |
| 1–8 | 904 | 14 | 50 | 15 | 854 | 14 | 186 | 21 | 6 | 12 | 180 | 21 |
| 9-17 | 542 | 8 | 39 | 12 | 503 | 8 | 79 | 15 | 3 | 8 | 76 | 15 |
| 18–49 | 2451 | 37 | 98 | 30 | 2353 | 38 | 533 | 22 | 20 | 20 | 513 | 22 |
| 50–64 | 1254 | 19 | 70 | 22 | 1184 | 19 | 380 | 30 | 11 | 16 | 369 | 31 |
| ≥ 65 | 1410 | 21 | 66 | 20 | 1344 | 22 | 815 | 58 | 32 | 48 | 783 | 58 |
| Median (IQR) | 42 (21-62) | | 42 (15-62) | | 42 (21-62) | | 59 (36-72) | | 60 (39-71) | | 59 (36-72) | |
| **Sex** | | | | | | | | | | | | |
| Female | 4022 | 61 | 177 | 55 | 3845 | 62 | 1270 | 32 | 44 | 25 | 1226 | 32 |
| Male | 2512 | 38 | 145 | 45 | 2367 | 38 | 722 | 29 | 28 | 19 | 694 | 29 |
| Unknown | 27 | 0 | 1 | 0 | 26 | 0 | 1 | 4 | 0 | 0 | 1 | 4 |
| **Comorbidity**[c] | | | | | | | | | | | | |
| No | 4621 | 70 | 235 | 73 | 4386 | 70 | 1095 | 24 | 46 | 20 | 1049 | 24 |
| Yes | 1561 | 24 | 75 | 23 | 1486 | 24 | 750 | 48 | 23 | 31 | 727 | 49 |
| Unknown | 379 | 6 | 13 | 4 | 366 | 6 | 148 | 39 | 3 | 23 | 145 | 40 |
| **Province** | | | | | | | | | | | | |
| Alberta | 908 | 14 | 45 | 14 | 863 | 14 | 306 | 34 | 10 | 22 | 296 | 34 |
| British Columbia | 1259 | 19 | 54 | 17 | 1205 | 19 | 521 | 41 | 20 | 37 | 501 | 42 |
| Ontario | 2944 | 45 | 132 | 41 | 2812 | 45 | 948 | 32 | 33 | 25 | 915 | 33 |
| Quebec | 1450 | 22 | 92 | 28 | 1358 | 22 | 218 | 15 | 9 | 10 | 209 | 15 |
| **Weeks of specimen collection, 2025/26**[d] | | | | | | | | | | | | |
| 44-47 | 788 | 12 | 43 | 13 | 745 | 12 | 104 | 13 | 5 | 12 | 99 | 13 |
| 48-51 | 1471 | 22 | 157 | 49 | 1314 | 21 | 421 | 29 | 29 | 18 | 392 | 30 |
| 52-2[e] | 1008 | 15 | 82 | 25 | 926 | 15 | 348 | 35 | 26 | 32 | 322 | 35 |
| 3-6 | 1070 | 16 | 23 | 7 | 1047 | 17 | 359 | 34 | 7 | 30 | 352 | 34 |
| 7-10 | 958 | 15 | 9 | 3 | 949 | 15 | 321 | 34 | 2 | 22 | 319 | 34 |
| 11-14 | 751 | 11 | 4 | 1 | 747 | 12 | 277 | 37 | 0 | 0 | 277 | 37 |
| 15-17 | 515 | 8 | 5 | 2 | 510 | 8 | 163 | 32 | 3 | 60 | 160 | 31 |
| **2024/25 influenza vaccination status**[f] | | | | | | | | | | | | |
| No | 3614 | 55 | 203 | 63 | 3411 | 55 | 231 | 6 | 5 | 2 | 226 | 7 |
| Yes | 2134 | 33 | 82 | 25 | 2052 | 33 | 1484 | 70 | 55 | 67 | 1429 | 70 |
| Unknown | 813 | 12 | 38 | 12 | 775 | 12 | 278 | 34 | 12 | 32 | 266 | 34 |

Abbreviations: ARI, acute respiratory illness; IQR, interquartile range.

[a] 2025/26 vaccination status based on participant or guardian report. Participants vaccinated <2 weeks before onset of symptoms or with unknown vaccination status or timing were excluded. Only trivalent formulations were used in Canada with virtually all publicly-funded vaccines in SPSN provinces being inactivated (≥99% overall; <5% live-attenuated in BC and Quebec) and egg-based (≥90% overall; <30% cell-based in Alberta and <5% in Ontario). In all provinces, adjuvanted vaccines were available for community-dwelling older adults (≥65 years, ≥75 years in Quebec), with high dose vaccines also offered in Ontario.

[b] Children < 1 year excluded based on variability and/or uncertainty in their age-related vaccine eligibility over the course of the epidemic. Older age strata defined as per usual SPSN analyses predicated upon higher likelihood of chronic comorbidity at ≥ 50 years and higher age-associated risk among adults ≥ 65 years [7].

[c] Includes chronic comorbidities that place individuals at higher risk of serious complications from influenza as defined by Canada's National Advisory Committee on Immunization [7].

[d] Missing specimen collection dates were imputed as the date the specimen was received and processed at the laboratory minus 2 days.

[e] Includes epi-weeks 52, 53, 1, and 2.

[f] 2024/25 vaccination status based on participant or guardian report.

**Supplementary Table S7.** Participant profile, influenza B analyses, Canadian Sentinel Practitioner Surveillance Network, 26 October 2025 – 02 May 2026 (epi-weeks 44–17).

| Characteristics | All ARI participants (column %) | | | | | | Influenza vaccinated[a] (row %) | | | | | |
|---|---|---|---|---|---|---|---|---|---|---|---|---|
| | Overall | | Influenza B cases | | Influenza controls | | Overall | | Influenza B cases | | Influenza controls | |
| | n | % | n | % | n | % | n | % | n | % | n | % |
| N (row %) | 6742 | 100 | 504 | 7 | 6238 | 93 | 1991 | 30 | 70 | 14 | 1921 | 31 |
| **Age group (years)** [b] | | | | | | | | | | | | |
| 1–8 | 964 | 14 | 110 | 22 | 854 | 14 | 188 | 20 | 8 | 7 | 180 | 21 |
| 9-17 | 627 | 9 | 124 | 25 | 503 | 8 | 99 | 16 | 23 | 19 | 76 | 15 |
| 18–49 | 2578 | 38 | 225 | 45 | 2353 | 38 | 541 | 21 | 28 | 12 | 513 | 22 |
| 50–64 | 1220 | 18 | 36 | 7 | 1184 | 19 | 375 | 31 | 6 | 17 | 369 | 31 |
| ≥ 65 | 1353 | 20 | 9 | 2 | 1344 | 22 | 788 | 58 | 5 | 56 | 783 | 58 |
| Median (IQR) | 41 (19-61) | | 21 (10-41) | | 42 (21-62) | | 58 (35-72) | | 26 (11-42) | | 59 (36-72) | |
| **Sex** | | | | | | | | | | | | |
| Female | 4116 | 61 | 271 | 54 | 3845 | 62 | 1270 | 31 | 44 | 16 | 1226 | 32 |
| Male | 2598 | 39 | 231 | 46 | 2367 | 38 | 720 | 28 | 26 | 11 | 694 | 29 |
| Unknown | 28 | 0 | 2 | 0 | 26 | 0 | 1 | 4 | 0 | 0 | 1 | 4 |
| **Comorbidity**[c] | | | | | | | | | | | | |
| No | 4817 | 71 | 431 | 86 | 4386 | 70 | 1104 | 23 | 55 | 13 | 1049 | 24 |
| Yes | 1540 | 23 | 54 | 11 | 1486 | 24 | 738 | 48 | 11 | 20 | 727 | 49 |
| Unknown | 385 | 6 | 19 | 4 | 366 | 6 | 149 | 39 | 4 | 21 | 145 | 40 |
| **Province** | | | | | | | | | | | | |
| Alberta | 991 | 15 | 128 | 25 | 863 | 14 | 320 | 32 | 24 | 19 | 296 | 34 |
| British Columbia | 1280 | 19 | 75 | 15 | 1205 | 19 | 514 | 40 | 13 | 17 | 501 | 42 |
| Ontario | 3038 | 45 | 226 | 45 | 2812 | 45 | 946 | 31 | 31 | 14 | 915 | 33 |
| Quebec | 1433 | 21 | 75 | 15 | 1358 | 22 | 211 | 15 | 2 | 3 | 209 | 15 |
| **Weeks of specimen collection, 2025/26**[d] | | | | | | | | | | | | |
| 44-47 | 747 | 11 | 2 | 0 | 745 | 12 | 99 | 13 | 0 | 0 | 99 | 13 |
| 48-51 | 1332 | 20 | 18 | 4 | 1314 | 21 | 394 | 30 | 2 | 11 | 392 | 30 |
| 52-2[e] | 947 | 14 | 21 | 4 | 926 | 15 | 322 | 34 | 0 | 0 | 322 | 35 |
| 3-6 | 1116 | 17 | 69 | 14 | 1047 | 17 | 357 | 32 | 5 | 7 | 352 | 34 |
| 7-10 | 1109 | 16 | 160 | 32 | 949 | 15 | 349 | 31 | 30 | 19 | 319 | 34 |
| 11-14 | 893 | 13 | 146 | 29 | 747 | 12 | 301 | 34 | 24 | 16 | 277 | 37 |
| 15-17 | 598 | 9 | 88 | 17 | 510 | 8 | 169 | 28 | 9 | 10 | 160 | 31 |
| **2024/25 influenza vaccination status**[f] | | | | | | | | | | | | |
| No | 3766 | 56 | 355 | 70 | 3411 | 55 | 240 | 6 | 14 | 4 | 226 | 7 |
| Yes | 2136 | 32 | 84 | 17 | 2052 | 33 | 1478 | 69 | 49 | 58 | 1429 | 70 |
| Unknown | 840 | 12 | 65 | 13 | 775 | 12 | 273 | 33 | 7 | 11 | 266 | 34 |

Abbreviations: ARI, acute respiratory illness; IQR, interquartile range.

[a] 2025/26 vaccination status based on participant or guardian report. Participants vaccinated <2 weeks before onset of symptoms or with unknown vaccination status or timing were excluded. Only trivalent formulations were used in Canada with virtually all publicly-funded vaccines in SPSN provinces being inactivated (≥99% overall; <5% live-attenuated in BC and Quebec) and egg-based (≥90% overall; <30% cell-based in Alberta and <5% in Ontario). In all provinces, adjuvanted vaccines were available for community-dwelling older adults (≥65 years, ≥75 years in Quebec), with high dose vaccines also offered in Ontario.

[b] Children < 1 year excluded based on variability and/or uncertainty in their age-related vaccine eligibility over the course of the epidemic. Older age strata defined as per usual SPSN analyses predicated upon higher likelihood of chronic comorbidity at ≥ 50 years and higher age-associated risk among adults ≥ 65 years [7].

[c] Includes chronic comorbidities that place individuals at higher risk of serious complications from influenza as defined by Canada's National Advisory Committee on Immunization [7].

[d] Missing specimen collection dates were imputed as the date the specimen was received and processed at the laboratory minus 2 days.

[e] Includes epi-weeks 52, 53, 1, and 2.

[f] 2024/25 vaccination status based on participant or guardian report.

**Supplementary Table S8.** COVID-19 vaccination status among participants vaccinated for influenza, Canadian Sentinel Practitioner Surveillance Network, 26 October 2025 – 02 May 2026 (epi-weeks 44-17)

| | **COVID-19 vaccination status** | | | |
|---|---|---|---|---|
| | **Vaccinated (n)** | **Vaccinated (%)** | **Unvaccinated (n)** | **Total (n)** |
| **Total** | 1154 | 51% | 1107 | 2261 |
| Influenza A | 196 | 49% | 207 | 403 |
| Influenza B | 23 | 33% | 46 | 69 |
| Influenza test-negative control | 935 | 52% | 854 | 1789 |
| 1-17 years | 68 | 29% | 168 | 236 |
| 18-64 years | 382 | 46% | 442 | 824 |
| ≥65 years | 485 | 67% | 244 | 729 |

Participants restricted to those vaccinated for influenza. Influenza and COVID-19 vaccination status based on self-report and receipt ≥2 weeks prior to onset; those vaccinated <2 weeks before onset or with unknown vaccination status are excluded

**Supplementary Figure S1.** Median age of unvaccinated influenza A cases and controls, Canadian Sentinel Practitioner Surveillance Network, 2009/10 to 2025/26.

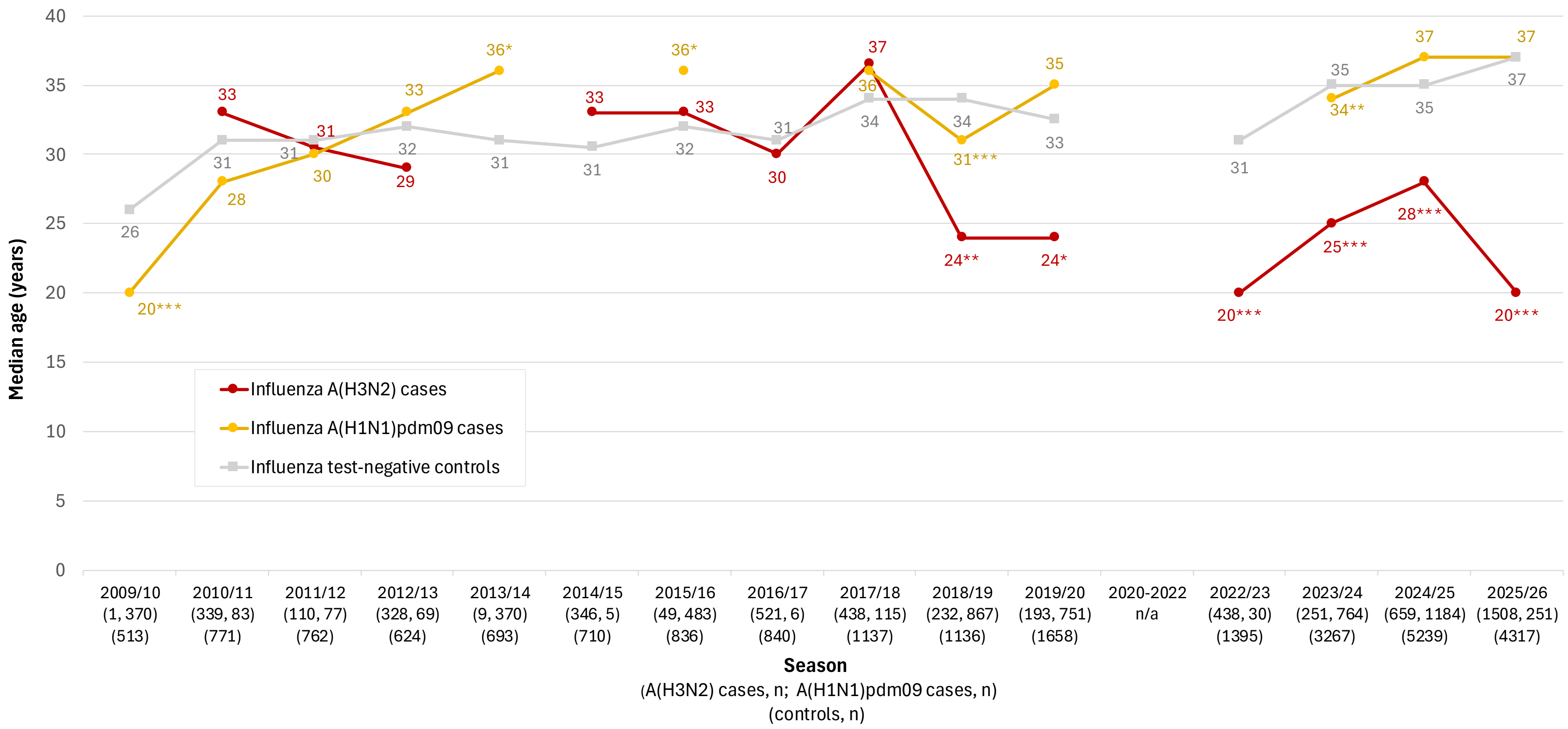

Median age not displayed in figure if based on ≤ 30 specimens. Age distributions compared using Wilcoxon rank-sum test. Median, interquartile range, and p-values are available in **Supplementary Table S9**.

* median age of influenza cases significantly different from controls (p<0.05)
** median age of influenza cases significantly different from controls (p<0.01)
*** median age of influenza cases significantly different from controls (p<0.0001)

**Supplementary Table S9.** Age distribution of unvaccinated influenza A cases and controls, Canadian Sentinel Practitioner Surveillance Network, 2009/10 to 2025/26.

| | Influenza A(H3N2)[a] | | | | Influenza A(H1N1)pdm09[a] | | | | Influenza controls[a] | | | | p-value[b] | | |
|---|---|---|---|---|---|---|---|---|---|---|---|---|---|---|---|
| **Season[c,d]** | **N** | **Q1** | **Median** | **Q3** | **N** | **Q1** | **Median** | **Q3** | **N** | **Q1** | **Median** | **Q3** | **A(H3N2) vs control** | **A(H1N1) vs control** | **A(H3N2) vs A(H1N1)** |
| 2009/10 | **1** | - | - | - | **370** | 10 | 20 | 33 | **513** | 16 | 26 | 42 | 0.262 | - | - |
| 2010/11 | **339** | 16 | 33 | 46 | **83** | 22 | 28 | 37 | **771** | 17 | 31 | 45 | 0.427 | 0.975 | 0.392 |
| 2011/12 | **110** | 13 | 31 | 44 | **77** | 22 | 30 | 41 | **762** | 17 | 31 | 45 | 0.592 | 0.929 | 0.902 |
| 2012/13 | **328** | 13 | 29 | 47 | **69** | 25 | 33 | 49 | **624** | 20 | 32 | 48 | 0.086 | 0.408 | 0.068 |
| 2013/14 | **9** | 15 | 22 | 37 | **370** | 23 | 36 | 49 | **693** | 19 | 31 | 47 | 0.297 | 0.021 | 0.118 |
| 2014/15 | **346** | 15 | 33 | 49 | **5** | 23 | 30 | 31 | **710** | 18 | 31 | 48 | 0.809 | 0.628 | 0.562 |
| 2015/16 | **49** | 20 | 33 | 51 | **483** | 21 | 36 | 48 | **836** | 17 | 32 | 48 | 0.386 | 0.031 | 0.941 |
| 2016/17 | **521** | 15 | 30 | 51 | **6** | 16 | 48 | 60 | **840** | 17 | 31 | 49 | 0.897 | 0.264 | 0.270 |
| 2017/18 | **438** | 20 | 37 | 54 | **115** | 16 | 36 | 48 | **1137** | 20 | 34 | 49 | 0.097 | 0.754 | 0.175 |
| 2018/19 | **232** | 15 | 24 | 46 | **867** | 8 | 31 | 45 | **1136** | 18 | 34 | 50 | 0.007 | <.0001 | 0.635 |
| 2019/20 | **193** | 12 | 24 | 46 | **751** | 22 | 35 | 47 | **1658** | 17 | 33 | 49 | 0.018 | 0.114 | 0.003 |
| 2020-2022[e] | **-** | - | - | - | **-** | - | - | - | **-** | - | - | - | - | - | - |
| 2022/23 | **438** | 8 | 20 | 37 | **30** | 13 | 43 | 55 | **1395** | 10 | 31 | 50 | <.0001 | 0.128 | 0.003 |
| 2023/24 | **251** | 13 | 25 | 38 | **764** | 7 | 34 | 49 | **3267** | 14 | 35 | 52 | <.0001 | 0.004 | 0.087 |
| 2024/25 | **659** | 11 | 28 | 43 | **1184** | 12 | 37 | 51 | **5239** | 13 | 35 | 52 | <.0001 | 0.907 | <0.0001 |
| 2025/26 | **1508** | 9 | 20 | 40 | **251** | 13 | 37 | 57 | **4317** | 17 | 37 | 55 | <.0001 | 0.986 | <0.0001 |

Abbreviations: N, number; Q1, first quartile; Q3, third quartile

[a] Displayed summary statistics are shown for age in years.
[b] Age distributions compared using Wilcoxon rank-sum test
[c] Seasons span from epi-weeks 44 to 17 or 18, except for in 2019/20 (epi-weeks 44 to 13) and 2009/10 (epi-weeks 44 to 6).
[d] From 2009/10 to 2022/23, influenza-like illness (ILI) was used as the case definition for recruitment (defined as new or worsening cough and fever (fever not required among ≥65 years) and one or more of sore throat, arthralgia, myalgia, or prostration). Starting in 2023/24, acute respiratory illness (ARI) was used (defined as new or worsening cough). Most ARI participants also met historic ILI case definition (>85% of cases and >75% of controls), including in 2023/24 (92% of A(H3N2), 87% of A(H1N1)pdm09, 76% of controls), 2024/25 (90% of A(H3N2), 90% of A(H1N1)pdm09, 78% of controls), and 2025/26 (89% of A(H3N2), 86% of A(H1N1)pdm09, 78% of controls).
[e] Not displayed for 2020/21 to 2021/22 seasons given lack of influenza circulation during the COVID-19 pandemic.

**Supplementary Table S10.** Age distribution of unvaccinated participants by influenza type, subtype, and subclade, Canadian Sentinel Practitioner Surveillance Network, 26 October 2025 – 02 May 2026 (epi-weeks 44–17)

| | Total | 1-8 years | 9-17 years | 18-29 years | 30-49 years | 50-64 years | ≥65 years | Median age (IQR), years |
|---|---|---|---|---|---|---|---|---|
| **Influenza controls** | | | | | | | | |
| Overall, n (row %) | 4317 | 674 (16%) | 427 (10%) | 533 (12%) | 1307 (30%) | 815 (19%) | 561 (13%) | 37 (17-55) |
| **Influenza A(H3N2)** | | | | | | | | |
| Overall, n (row %) | 1508 | 371 (25%) | 343 (23%) | 222 (15%) | 313 (21%) | 155 (10%) | 104 (7%) | 20 (9-40) |
| Subclade K, n (row %, column %[a]) | 763 | 200 (26%, 88%) | 171 (22%, 91%) | 121 (16%, 89%) | 139 (18%, 80%) | 80 (10%, 84%) | 52 (7%, 90%) | 18 (8-40) |
| Subclade J.2.4.2[b], n (row %, column %[a]) | 82 | 17 (21%, 7%) | 16 (20%, 9%) | 11 (13%, 8%) | 25 (30%, 14%) | 10 (12%, 11%) | 3 (4%, 5%) | 26 (11-42) |
| Other subclades[c] , n (row %, column %[a]) | 33 | 11 (33%, 5%) | 1 (3%, 1%) | 4 (12%, 3%) | 9 (27%, 5%) | 5 (15%, 5%) | 3 (9%, 5%) | 32 (6-47) |
| Not sequenced[f] n (row%, column %[a]) | 630 | 143 (23%, 39%) | 155 (25%, 45%) | 86 (14%, 39%) | 140 (22%, 45%) | 60 (10%, 39%) | 46 (7%, 44%) | 20 (10-40) |
| **Influenza A(H1N1)pdm09** | | | | | | | | |
| Overall, n (row %) | 251 | 44 (18%) | 36 (14%) | 21 (8%) | 57 (23%) | 59 (24%) | 34 (14%) | 37 (13-57) |
| Subclade D.3.1, n (row %, column %[a]) | 33 | 5 (15%, 18%) | 8 (24%, 31%) | 2 (6%, 13%) | 11 (33%, 27%) | 4 (12%, 11%) | 3 (9%, 12%) | 34 (14-43) |
| Subclade D.3.1.1, n (row %, column %[a]) | 139 | 23 (17%, 82%) | 18 (13%, 69%) | 14 (10%, 88%) | 30 (22%, 73%) | 32 (23%, 89%) | 22 (16%, 88%) | 37 (13-60) |
| Not sequenced[f] n (row %, column %[a]) | 75 | 16 (21%, 36%) | 10 (13%, 28%) | 5 (7%, 24%) | 14 (19%, 25%) | 21 (28%, 36%) | 9 (12%, 26%) | 41 (10-54) |
| **Influenza B** | | | | | | | | |
| Overall, n (row %) | 434 | 102 (24%) | 101 (23%) | 44 (10%) | 153 (35%) | 30 (7%) | 4 (1%) | 20 (9-40) |
| Subclade C.3[d], n (row %, column %[a]) | 55 | 17 (31%, 20%) | 17 (31%, 18%) | 4 (7%, 11%) | 15 (27%, 12%) | 2 (4%, 8%) | 0 (0%, 0%) | 11 (8-33) |
| Subclade C.5[e] , n (row %, column %[a]) | 308 | 70 (23%, 80%) | 75 (24%, 82%) | 32 (10%, 89%) | 106 (34%, 88%) | 24 (8%, 92%) | 1 (0%, 100%) | 20 (10-40) |
| Not sequenced[f], n (row %, column %[a]) | 71 | 15 (21%, 15%) | 9 (13%, 9%) | 8 (11%, 18%) | 32 (45%, 21%) | 4 (6%, 13%) | 3 (4%, 75%) | 33 (12-45) |

Abbreviations: IQR, interquartile range.

---

[a] Column percent for sequenced specimens is relative to total displayed sequenced specimens per subtype and age group (e.g., in 1-8 years: 200 subclade K specimens / 228 total displayed sequenced specimens = 88% subclade K). Column percent for non-sequenced specimens is relative to overall total (e.g., in 1-8 years: 143 non-sequenced specimens / 371 total A(H3N2) cases = 39% non-sequenced).

[b] Subclade K (formerly known as subclade J.2.4.1) and subclade J.2.4.2 viruses share parental subclade J.2.4 clade-defining substitutions T135K (A)(RBS)(-CHO) and K189R (B), plus additional S144N (A)(+CHO), N158D (B), I160K (B) and T328A mutations. Subclade K is differentiated by additional K2N, Q173R (D) and HA2: S49N, while subclade J.2.4.2 is differentiated by F79V. For more details see **Supplementary Table S1**.

[c] Includes subclade J.2, J.2.2, J.2.3, J.2.4.

[d] Includes subclade C.3.1 and C.3.3. For more details, see **Supplementary Table S2**

[e] Includes subclade C.5, C.5.1, C.5.6, C.5.6.1, and C.5.6. For more details, see **Supplementary Table S2**.

[f] Includes those that failed sequencing or have pending sequencing results. Over two-thirds of specimens in this category failed sequencing.

**Supplementary Table S11.** Vaccine effectiveness against influenza, sensitivity analyses, Canadian Sentinel Practitioner Surveillance Network, 26 October 2025 – 02 May 2026 (epi-weeks 44-17)

| | Total | Cases n vac[a] / N total (%) | Controls n vac[a] / N total (%) | VE [b c] (95% CI) |
|---|---|---|---|---|
| **Primary analysis** | | | | |
| Influenza A(H3N2) | 8086 | 340/1848 (18%) | 1921/6238 (31%) | 38 (27, 47) |
| 1-17 years | 2168 | 97/811 (12%) | 256/1357 (19%) | 37 (14, 54) |
| 18-64 years | 4347 | 120/810 (15%) | 882/3537 (25%) | 46 (32, 57) |
| ≥ 65 years | 1571 | 123/227 (17%) | 783/1344 (58%) | 15 (-17, 39) |
| Influenza A(H1N1)pdm09 | 6561 | 72/323 (22%) | 1921/6238 (31%) | 28 (4, 47) |
| 1-17 years | 1446 | 9/89 (10%) | 256/1357 (19%) | 41 (-25, 72) |
| 18-64 years | 3705 | 31/168 (18%) | 882/3537 (25%) | 26 (-13, 51) |
| ≥ 65 years | 1410 | 32/66 (48%) | 783/1344 (58%) | 24 (-29, 55) |
| Influenza B | 6742 | 70/504 (14%) | 1921/6238 (31%) | 55 (41, 66) |
| 1-17 years | 1591 | 31/234 (13%) | 256/1357 (19%) | 49 (21, 67) |
| ≥18 years | 5151 | 39/270 (14%) | 1665/4881 (34%) | 59 (41, 72) |
| **Excluding COVID-19 detections from controls** | | | | |
| Influenza A(H3N2) | 7694 | 340/1848 (18%) | 1802/5846 (31%) | 38 (27, 47) |
| 1-17 years | 2106 | 97/811 (12%) | 240/1295 (19%) | 37 (13, 54) |
| 18-64 years | 4117 | 120/810 (15%) | 838/3307 (25%) | 46 (32, 58) |
| ≥ 65 years | 1471 | 123/227 (54%) | 724/1244 (58%) | 17 (-16, 40) |
| Influenza A(H1N1)pdm09 | 6169 | 72/323 (22%) | 1802/5846 (31%) | 29 (4, 47) |
| 1-17 years | 1384 | 9/89 (10%) | 240/1295 (19%) | 40 (-27, 71) |
| 18-64 years | 3475 | 31/168 (18%) | 838/3307 (25%) | 26 (-12, 52) |
| ≥ 65 years | 1310 | 32/66 (48%) | 724/1244 (58%) | 25 (-27, 56) |
| Influenza B | 6355 | 70/504 (14%) | 1803/5851 (31%) | 55 (41, 66) |
| 1-17 years | 1532 | 31/234 (13%) | 240/1298 (18%) | 47 (18, 66) |
| ≥18 years | 4823 | 39/270 (14%) | 1563/4553 (34%) | 60 (42, 72) |
| **Additional adjustment for sex and comorbidity** | | | | |
| Influenza A(H3N2) | 7601 | 314/1751 (18%) | 1775/5850 (30%) | 38 (26, 47) |
| 1-17 years | 2055 | 90/772 (12%) | 239/1283 (19%) | 40 (16, 56) |
| 18-64 years | 4098 | 105/763 (14%) | 820/3335 (25%) | 47 (32, 59) |
| ≥ 65 years | 1448 | 119/216 (55%) | 716/1232 (58%) | 12 (-23, 37) |
| Influenza A(H1N1)pdm09 | 6159 | 69/309 (22%) | 1775/5850 (30%) | 25 (-1, 45) |
| 1-17 years | 1366 | 9/83 (11%) | 239/1283 (19%) | 33 (-43, 68) |
| 18-64 years | 3498 | 29/163 (18%) | 820/3335 (25%) | 24 (-18, 51) |
| ≥ 65 years | 1295 | 31/63 (49%) | 716/1232 (58%) | 20 (-38, 54) |
| Influenza B | 6333 | 66/483 (14%) | 1775/5850 (30%) | 54 (39, 66) |
| 1-17 years | 1510 | 30/227 (13%) | 239/1283 (19%) | 47 (17, 66) |
| ≥ 18 years | 4823 | 36/256 (14%) | 1536/4567 (34%) | 59 (40, 72) |

CI: confidence interval; vac: vaccinated; VE: vaccine effectiveness.

[a] Vaccination status per participant/guardian report. Participants vaccinated < 2 weeks before symptom onset or with unknown vaccination status were excluded.

[b] VE was calculated as (1 − OR) × 100%. ORs compare per cent positivity between vaccinated and unvaccinated participants using logistic regression. Firth's logistic regression was explored as needed to address small sample size; VE estimates differed by ≤ 3% (data not shown).

[c] VE was adjusted for age group (1–8, 9–17, 18–49, 50–64, ≥ 65 years), calendar time (bi-weekly epi-week), and province (Alberta, BC, Ontario, Quebec).

**Supplementary Table S12.** Vaccine effectiveness against influenza, stratified by prior and current vaccination status, Canadian Sentinel Practitioner Surveillance Network, 26 October 2025 – 02 May 2026 (epi-weeks 44-17) (n=7789)

| | Total | Cases | Controls | VE, % (95% CI) |
|---|---|---|---|---|
| **Influenza A(H3N2)** | 7065 | 1602 | 5463 | - |
| Neither season vaccination (reference) | 4243 | 1058 | 3185 | - |
| Prior season vaccination only | 884 | 261 | 623 | -21 (-47, 1) |
| Current season vaccination only | 265 | 39 | 226 | 41 (12, 60) |
| Both seasons vaccination | 1673 | 244 | 1429 | 37 (23, 48) |
| **Influenza A(H1N1)pdm09** | 5748 | 285 | 5463 | - |
| Neither season vaccination (reference) | 3383 | 198 | 3185 | - |
| Prior season vaccination only | 650 | 27 | 623 | 37 (4, 59) |
| Current season vaccination only | 231 | 5 | 226 | 59 (-3, 84) |
| Both seasons vaccination | 1484 | 55 | 1429 | 36 (10, 55) |
| **Influenza B** | 5902 | 439 | 5463 | - |
| Neither season vaccination (reference) | 3526 | 341 | 3185 | - |
| Prior season vaccination only | 658 | 35 | 623 | 16 (-24, 44) |
| Current season vaccination only | 240 | 14 | 226 | 62 (32, 79) |
| Both seasons vaccination | 1478 | 49 | 1429 | 52 (32, 65) |

CI: confidence interval; vac: vaccinated; VE: vaccine effectiveness.

Vaccination status per participant/guardian report. Participants vaccinated < 2 weeks before symptom onset in 2025/26 or with unknown vaccination status in 2024/25 or 2025/26 were excluded.

VE was calculated as (1 − OR) × 100%. ORs compare per cent positivity between vaccinated and unvaccinated participants using logistic regression; a four-level indicator variable was used, with neither season vaccination as the reference group. VE was adjusted for age group (1–8, 9–17, 18–49, 50–64, ≥ 65 years), calendar time (bi-weekly epi-week), and province (Alberta, BC, Ontario, Quebec). Firth's logistic regression was explored as needed to address small sample size; VE estimates differed by ≤ 1% (data not shown).

2. World Health Organization (WHO). Manual for the laboratory diagnosis and virological surveillance of influenza. Geneva: WHO; 2011.
